# Multivariate blood biomarkers capture resilience and resistance phenotypes across the Alzheimer’s disease spectrum

**DOI:** 10.64898/2026.08.03.26359415

**Authors:** Kalliopi Mavromati, Connor Dalby, Austin Dibble, Ganna Leonenko, Katherine Birditt, Stelios Lamprou, Lucie Tvrda, Ioannis Konstantinidis, Lynne Hughes, Maura Malpetti, Valentina Escott-Price, Monika Harvey, Alessio Fracasso, Michele Svanera, Terrence Quinn

## Abstract

Alzheimer’s disease pathology and cognitive outcomes frequently diverge, yet current single-axis definitions cannot identify resilient (high pathology, preserved cognition) and resistant (high risk, low pathology) subgroups reliably at scale, obscuring the mechanisms that uncouple pathological burden from cognitive decline. Here, we developed a multivariate blood-based framework integrating 19 molecular assays and six risk instruments in the Bio-Hermes-001 cohort (n=1,009). Unsupervised clustering identified resilient (n=91) and resistant (n=81) subgroups, together comprising 17% of the cohort, with distinct amyloid, tau, and neurodegeneration profiles. Amyloid-PET yielded convergent but only partially overlapping classifications. Proteomic, cytokine, and polygenic profiling further distinguished resistance through an APOE-centred genomic signature and resilience through neuroinflammatory markers associated with progression toward clinical Alzheimer’s disease. A four-biomarker panel (Aβ40, p-tau217, p-tau181, NfL) reproduced subgroup assignments with 83% accuracy. These findings support resilience and resistance as molecularly distinct subgroups and provide a scalable framework for pathology-informed stratification and mechanistic investigation.

## Main

Alzheimer’s disease (AD) is increasingly understood as a heterogeneous, multipathway disorder, driven by distinct pathological processes that vary across individuals [1–4]. This biological complexity cannot be fully captured by a single biomarker or pathological axis, creating a major challenge for pre-symptomatic stratification. Identifying individuals according to their multidimensional pathological burden is essential for understanding disease trajectories before cognitive damage accumulates. A multivariate approach could therefore widen the window for effective intervention while improving the precision with which therapeutic candidates are evaluated in clinical trials.

It is well known that pathological burden alone does not fully determine cognitive outcome. A subset of individuals diverges from expected pathology-cognition relationships [5–7], challenging our understanding of the mechanisms linking pathological burden to cognitive decline. Identifying and characterizing these subgroups would offer insight into the mechanisms underlying protection against dementia [8]. Such divergence can be understood within the broader framework of reserve, resilience, and maintenance, which describes how individuals differ in their capacity to preserve cognition in the face of age-related change, brain injury, and disease. [9, 10].

Within this framework, two phenotypes are particularly relevant for multivariate biomarker stratification: resilience and resistance. *Resilient* individuals sustain significant pathological burden without the expected cognitive deficit, while *resistant* individuals remain cognitively intact with low pathological burden despite carrying substantial risk of developing dementia [7, 11–13]. Once defined at scale, resilient and resistant populations provide a tractable opportunity for investigating their divergence from the expected pathological and cognitive trajectories. This is of direct translational relevance for a field in which most affected individuals remain undetected until symptom onset. Blood-based biomarkers offer a particular opportunity to build such a framework at scale [14], as recent advances now enable pre-symptomatic staging of AD pathology more broadly than has previously been feasible [15–20].

However, operationalizing resilient subtypes in terms of molecular pathology remains challenging. Existing approaches rely predominantly on cognitive performance adjusted for structural neuroimaging or sociodemographic proxies, with in vivo molecular AD biomarkers (amyloid PET, tau PET, CSF, or plasma) applied in a minority of studies [11, 21]. Where blood- or imaging-based biomarkers have been applied, studies have examined resilience in the context of a narrow pathological target, whether amyloid, tau, or a CSF composite [22, 23]. Since cognitive decline is rarely driven by a single mechanism, a multivariate, pathology-grounded framework for the identification of these subgroups is needed. Yet, no study has assessed resilient and resistant status comprehensively across a broad set of pathological axes simultaneously [21, 24].

Here, we present a multivariate, pathology-and risk-grounded biomarker framework for the systematic identification of resilient and resistant individuals, developed and applied in the Bio-Hermes-001 cohort using 19 molecular assays and six risk instruments (n = 1,001; 563 [56%] female; Fig. 1; [20]). Unsupervised clustering of pathology and risk markers identifies resilient and resistant individuals as a distinct subgroup, comprising 17% of the cohort, with characteristic A*β*, p-tau, and neurodegeneration marker (NfL, GFAP, sTREM2) profiles that separate them from the canonical trajectories. Blood-based and A*β* PET-based identification diverge in the individuals they classify as resilient or resistant, yet yield pathologically interpretable, cognitively similar groups. Proteomic, cytokine, and polygenic profiling further reveal distinct molecular patterns of each group, with resistance marked by a genomic signature centred on the protective APOE *ε*3*/ε*3 genotype, and resilience by neuroinflammatory markers specific to the transition to clinically manifest AD. Finally, we show that a four-biomarker panel (A*β*40, p-tau217, p-tau181, NfL) reproduces these groups with 83% accuracy, offering a scalable route to translating this framework into research and clinical settings. Together, these findings demonstrate that resilient and resistant phenotypes are distinct and reproducible across cognitive, pathological, risk, and molecular axes, supporting their use for pathology-informed case-mix adjustment and risk-stratified recruitment in clinical trials.

**Fig. 1:**
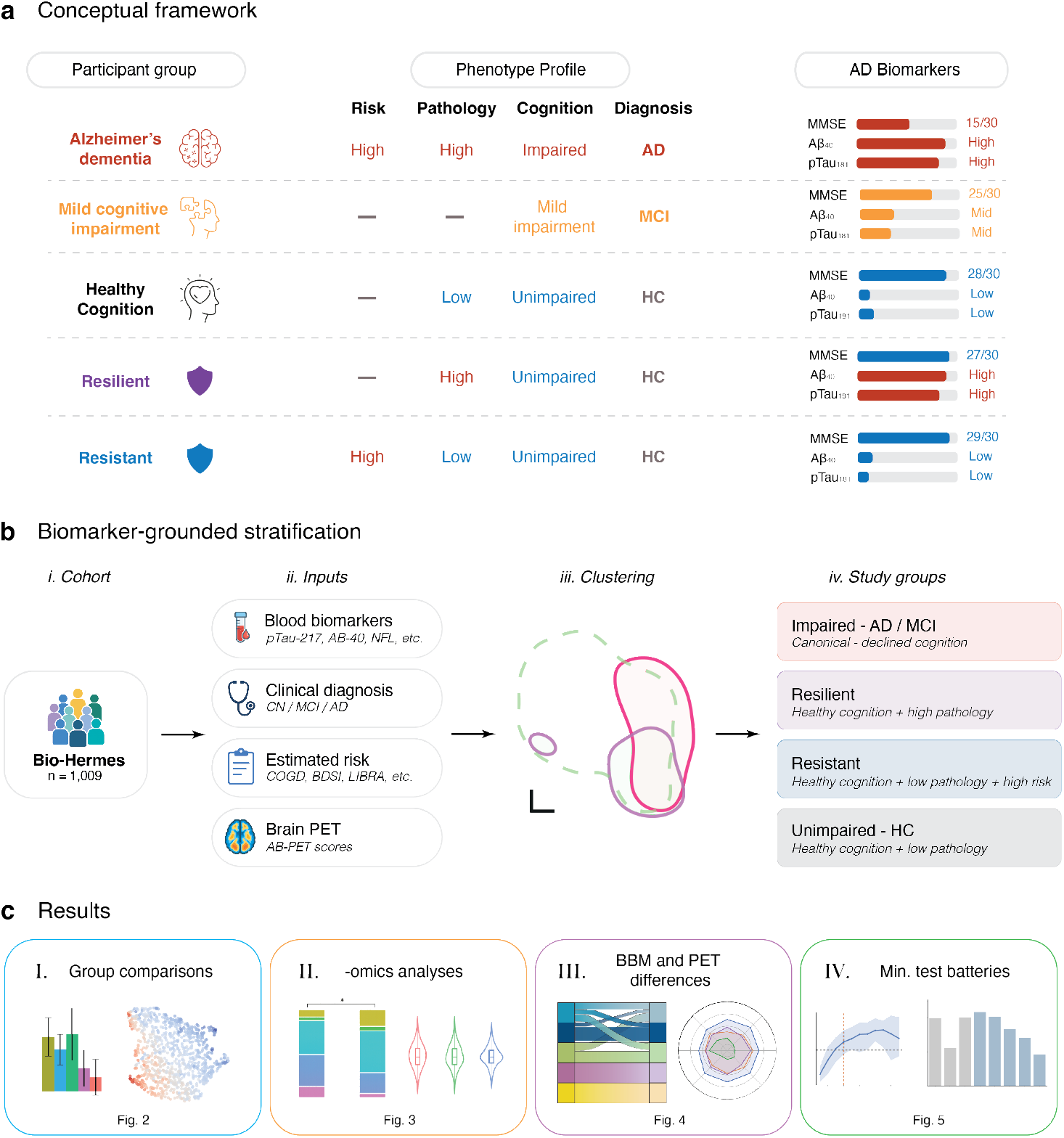
Framework overview. **a**, Conceptual framework defining five participant groups along a continuum of genetic and modifiable risk, pathology, and cognitive status. Alzheimer’s dementia (AD) and mild cognitive impairment (MCI) are defined by their level of cognitive impairment; healthy cognition (HC) individuals by low pathology and unimpaired cognition. Two atypical phenotypes are distinguished among clinically HC participants: *resilient* individuals maintain normal cognition despite high pathology; *resistant* individuals maintain normal cognition and low pathology despite high estimated risk. Representative late-life biomarker profiles (MMSE, plasma A*β*_40_, p-tau_181_) are shown per group; em dash (—) indicates that the feature is not used as part of the classification. **b**, Stratification pipeline. (*i*) Bio-Hermes-001 cohort (*n* = 1,009). (*ii*) Inputs: multimodal blood biomarkers (p-tau-217, A*β*_40_, NfL and others), clinical diagnosis (HC/MCI/AD), risk scores (COGDrisk, BDSI, LIBRA and others) and A*β*-PET. (*iii*) Unsupervised clustering partitioned high/low blood pathology and high/low risk into binary labels. (*iv*) Individuals were stratified into three canonical and two resilient/resistant groups. **c**, Downstream analyses: (*I*) group-level biomarker and risk comparisons; (*II*) multi-omic analyses; (*III*) blood-based biomarker (BBM) and PET differences across groups; and (*IV*) identification of min-

## Results

Within the Bio-Hermes-001 cohort (n = 1,001; 563 [56%] female; Method 2), our analytical pipeline identified two biologically distinct subgroups embedded within clinically healthy participants: a *resilient* group with high pathological burden despite preserved cognition, and a *resistant* group with high estimated AD risk but low pathological burden (Fig. 1; Method 18a). Together, these groups comprised approximately 17% of cohort, individuals invisible to standard clinical classification yet markedly biologically divergent. To distinguish these groups from other cognitively healthy individuals, we designed our pipeline to stratify healthy individuals on two axes: pathological burden, and dementia risk. We performed unsupervised clustering of blood-based biomarkers (BBMs; a proxy of pathological burden) and risk measures (Method 6; Extended Data Fig. 5) to produce binary designations of “high” or “low” pathology and risk per individual. Alongside the independent clinically adjudicated cognitive status (adjudicated via functional and cognitive testing alongside clinician’s judgement; Supplementary Table ST3), we used the pathology and risk classifications to designate individuals as resilient (n = 91) or resistant (n = 81); distinct from the healthy cognition (HC; n = 244), mild cognitive impairment (MCI; n = 308), and AD (n = 266) groups. To confirm that our yielded resilient and resistant groups match their theoretical definitions [7, 11, 24], we developed five PCA indices from the BBM and risk measures to perform a detailed comparison across our groups: All BBM (all 19 molecular assays), p-tau (all combined p-tau and Tau assays), A*β* (all combined A*β* assays), Risk (six risk instruments), and Glial/neurodegenerative (NDG; Fig. 2; Method 8; Supplementary Table ST15).

**Fig. 2:**
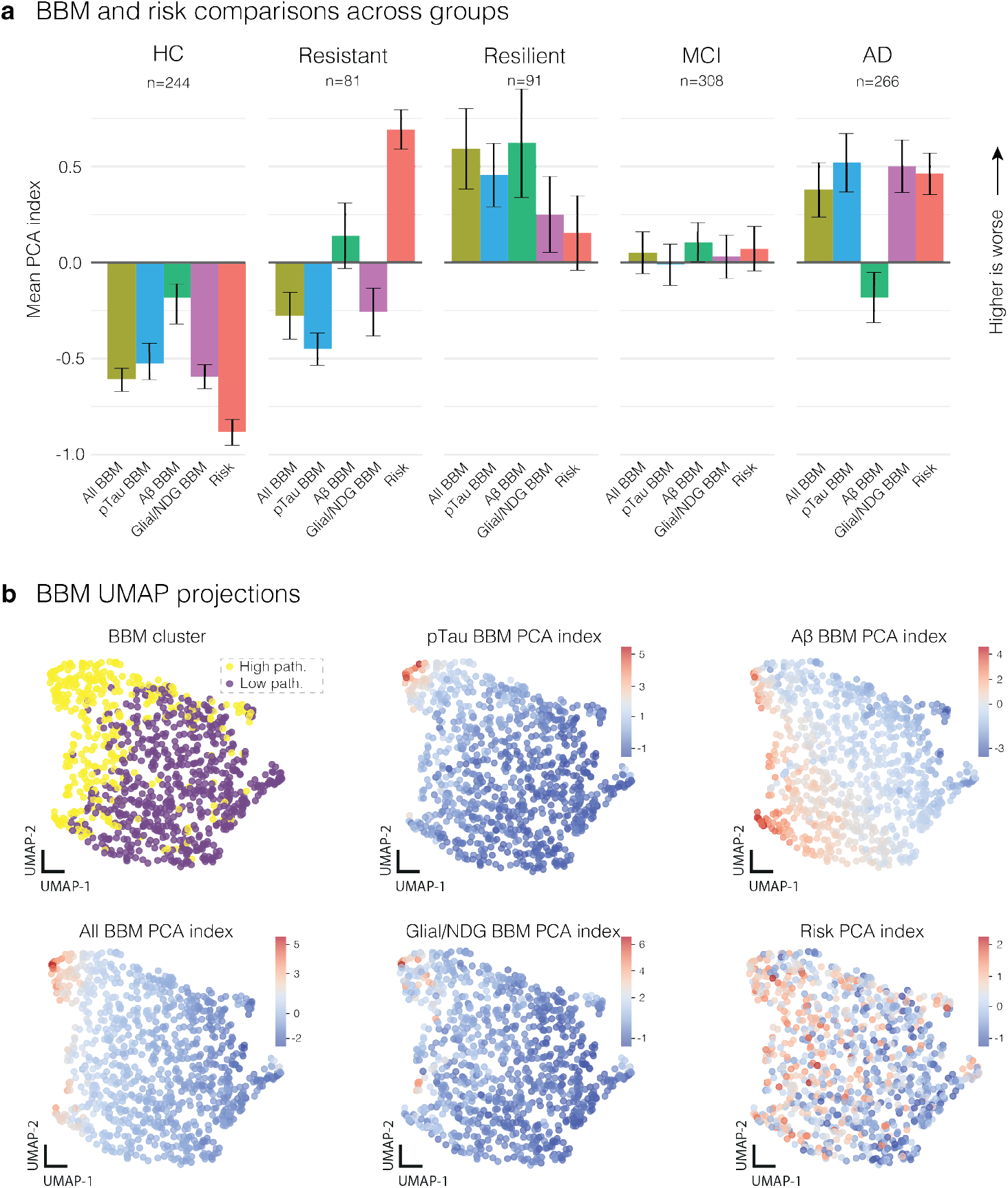
Blood-based biomarker (BBM) and demographic risk profiles across study groups. Biomarker and clustering profiles HC (n = 244), Resistant (n = 81), Resilient (n = 91), MCI (n = 308), and AD (n = 266). **a**, Group mean *±* s.d. across five PCA-derived indices (Method 8): All BBM (all BBM measures combined), p-tau (blood tau and phosphorylated tau measures), A*β* (blood A*β*40 and A*β*42 across vendors), Glial/neurodegenerative (NDG), and risk (composite of six demographic risk factors); higher values indicate worse outcome. HC shows the lowest values across all indices. Resistant individuals show the highest risk scores, with pathology intermediate between HC and MCI but A*β* comparable to MCI. Resilient individuals show the most consistently elevated pathology indices. MCI profiles cluster near zero across most indices. AD shows elevated pathology and risk overall, though A*β* is unexpectedly low relative to other pathology measures. **b**, UMAP [25] of BBM profiles across all individuals, coloured by a two-cluster solution distinguishing highversus low-BBM-pathology individuals (Method 6), and the continuous p-tau, A*β*, all BBM, glial/NDG, and risk indices (Method 8). High-p-tau, high-A*β*, and high-All-BBM individuals localize to distinct subregions coinciding with the high-pathology cluster. imal test batteries for efficient group discrimination.

## Resistant and resilient groups

### Resilient phenotype

Resilience does not represent asymptomatic AD pathology, but a distinct configuration in which substantial amyloid and tau burden coexists with preserved cognition, without the neuroinflammatory and neurodegenerative signal characteristic of clinical AD (GFAP, NfL, sTREM2; Fig. 2a). Resilient individuals carried the highest BBM-derived pathological burden of any group (p *<* 0.001; g = 0.66 [0.45, 0.88]) despite healthy diagnostic status, comparable in magnitude to clinically diagnosed AD, with all pathology indices significantly elevated relative to both the healthy cognition (HC) group and the mild cognitive impairment (MCI) group (p *<* 0.001, g = 0.95 [0.72, 1.17]). The composition of that signal, however, partially diverged from AD: blood A*β* was highest in the resilient group (p *<* 0.001; g = 0.70 [0.48, 0.92]), and p-tau did not differ significantly from AD (p = 0.562; g = *−*0.06 [*−*0.30, 0.18]). By contrast, several non-amyloid, non-tau pathology markers were significantly lower in the resilient group than in AD (p = 0.040; g = *−*0.23 [*−*0.47, 0.01]), indicating that this divergence was driven specifically by attenuated glial and neurodegenerative markers (GFAP, NfL, sTREM2) rather than by lower amyloid or tau.

### Resistant phenotype

Resistant individuals showed the highest estimated risk of any group (Fig. 2), consistent with their selection criterion, while remaining cognitively healthy throughout. BBM-derived pathological burden was significantly reduced relative to MCI and AD (p *<* 0.001; g = *−*0.47 [*−*0.70, *−*0.23]; Method 8), most clearly in p-tau (p *<* 0.001; g = *−*0.63 [*−*0.87, *−*0.40]). Pathological burden nonetheless remained elevated relative HC (p *<* 0.001; g = 0.66 [0.40, 0.91]), consistent with attenuation rather than absence of AD pathology. A*β* deviated markedly from this pattern: resistant individuals showed higher blood A*β* than HC (p = 0.002; g = 0.46 [0.21, 0.72]) and AD (p = 0.006; g = 0.31 [0.06, 0.56]), indistinguishable from MCI (p = 0.731; g = 0.04 [*−*0.21, 0.28]). This dissociation between A*β* and overall pathological burden constitutes a defining feature of the resistant profile, possibly reflecting differential peripheral clearance or altered production under sustained high risk [26, 27].

### MCI and AD reference profiles

The MCI group occupied an intermediate profile across risk and pathology indices, consistent with progressive pathological accumulation underlying cognitive decline. The resilient subgroup’s comparable pathological burden without impairment, however, challenges a linear burden-to-decline interpretation of the MCI profile. The aggregated MCI profile likely conflates biologically distinct subpopulations, with some individuals on a trajectory toward AD and others reflecting a different biological context. AD showed the expected pattern of elevated risk and BBM pathology (p *<* 0.001; g = 0.53 [0.39, 0.68]), except that blood A*β* was comparable to HC (p = 0.689; g = 0.03 [*−*0.14, 0.21]) and substantially lower than in resilient individuals. This dissociation, consistent with impaired peripheral clearance and cortical sequestration in established AD [26, 27] reinforces the case for multi-analyte profiling.

### Demographic characteristics

Resistant individuals showed the oldest age distribution, with the highest proportion of participants in the 70–79 and 80–89 year bands (Extended Data Fig. 1a, Supplementary Fig.S9). This pattern is expected by construction, since age enters composite risk instruments (Method 5), and is reported as a consistency check. Notably, despite this advanced age, resistant individuals showed lower pathology than MCI and AD. Resilient individuals were also older than HC, but not significantly different than MCI and AD.

### Validity of the stratification

Because the pathology and risk indices formed the basis for clustering, their divergence between the derived groups and HC (Fig. 2a) is circular by construction and reported to confirm the groups separated as intended. This heterogeneity, invisible to clinical HC classification, approached the magnitude seen in clinician-adjudicated MCI and AD, whose diagnoses were assigned independently of clustering. All subsequent comparisons, genomic, proteomic, cytokine, and A*β* PET (Figs. 3, 4), use measures not used in clustering and constitute the non-circular evidence for phenotype validity presented here.

**Fig. 3:**
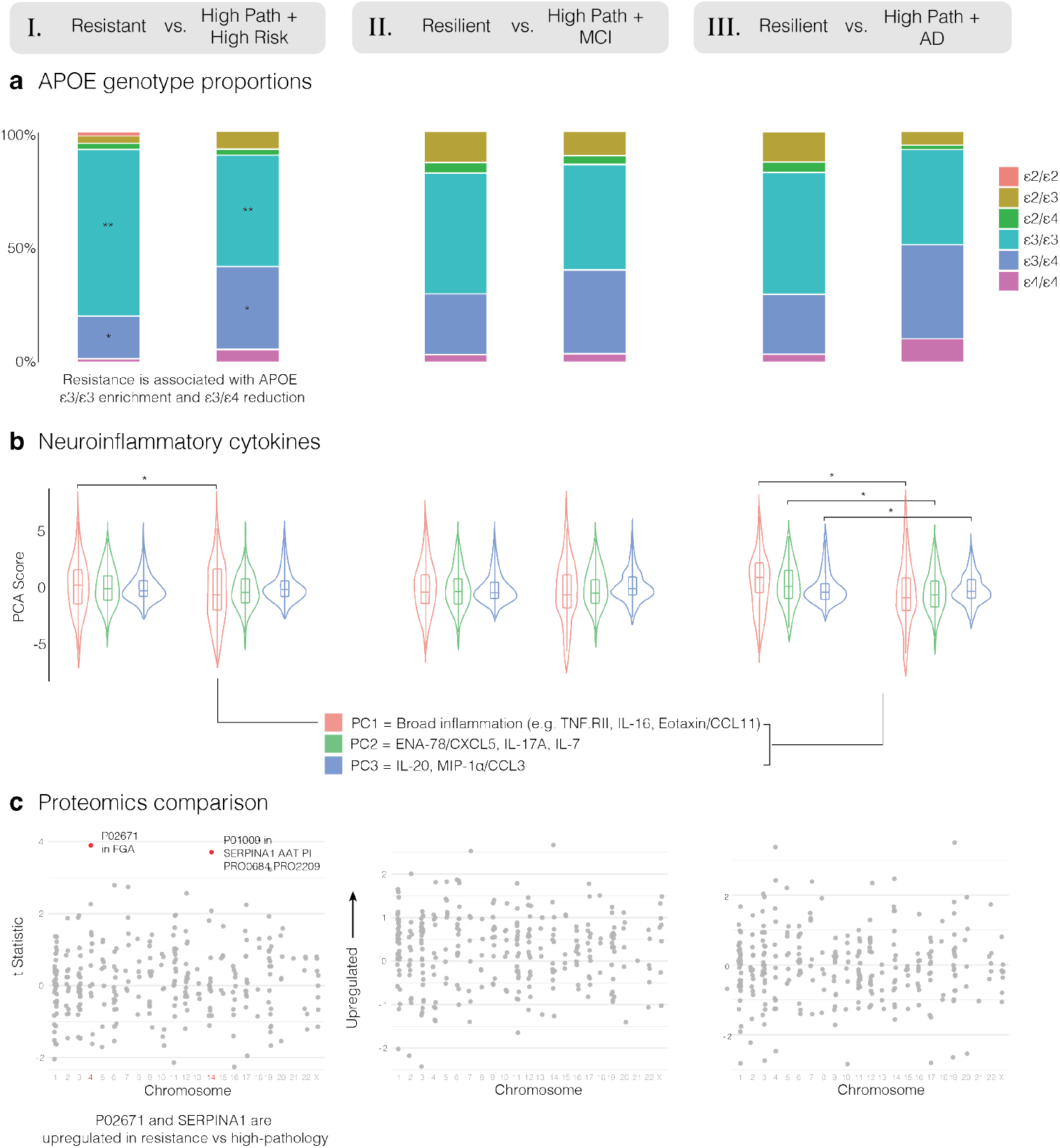
Multi-modal characterization of resilience to Alzheimer’s disease neuropathology across cognitive outcomes. Comparisons: I. Resistant vs. High Pathology + High Risk (left), II. Resilient vs. High Pathology + MCI (middle), and III. Resilient vs. High Pathology + AD (right). **a**, APOE genotype proportions. Stacked bar charts depicting the distribution of APOE genotypes in each group. Comparison I shows significant enrichment of *ε*4-containing alleles in High Pathology + High Risk group relative to Resistant, alongside *ε*3/*ε*3 enrichment in the Resistant (**P *<* 0.001, *P *<* 0.01, Fisher exact test). Comparison III shows potential *ε*3/*ε*4 reduction in Resilient relative to High Pathology + AD (nominally significant P = 0.020 before multiple comparisons correction). No difference in comparison II (Supplementary Tables ST5 and ST6; Method 12). **b**, Cytokine PC scores (PC1, salmon = broad inflammation; PC2, green = ENA-78/CXCL5, IL-17A, IL-7; PC3, blue = IL-20, MIP-1*α*/CCL3). Comparison I: significant increase in PC1 (*P *<* 0.05, logistic regression). Comparison III: significant PC1, PC2, and PC3 between differences (*P *<* 0.05 for each). No differences in comparison II (Supplementary Table ST7). **c**, Proteomics tstatistics by chromosomal position; significant proteins annotated (red). Comparison I: FGA (P02671, chromosome 4) and SERPINA1/AAT (P01009, chromosome 14) upregulated in the Resistant vs. High Pathology + High Risk (Method 14). No significant proteins in comparisons II or III.

Collectively, these results show that multivariate blood-based stratification resolves heterogeneity within cognitively healthy populations relevant to prevention trial design and early intervention.

### Molecular patterns of resilience and resistance

To characterize the molecular basis of preserved cognition despite elevated pathology or risk, we compared resistant and resilient individuals against paired counterfactual groups sharing equivalent pathological and risk profiles but differing in cognitive outcome, across three comparisons (Fig. 3; Supplementary Tables ST5–ST7). Cognitive performance was significantly higher in resistant and resilient individuals in all three comparisons (p *<* 0.001, two-sided Wilcoxon rank-sum test), confirming the intended divergence in cognitive outcome (Extended Data Fig. 3).

### Genomic architecture of resistance

The clearest molecular distinction between resistant individuals and their high-risk, high-pathology counterparts was genomic (Method 12). APOE *ε*2 and *ε*4 carriership together significantly discriminated the groups (*β* = 0.45, p = 0.007, AUC = 0.63; Supplementary Table ST5), driven by marked enrichment in the *ε*3/*ε*3 genotype in the resistant group, absent from all other comparisons (Fig. 3a). ADRD PRS most strongly discriminated the groups (*β* = 0.57, p *<* 0.001, AUC = 0.65), with an attenuated but significant effect after APOE exclusion (b = 0.38, p = 0.007, AUC = 0.61; full PRS specification in Supplementary Table ST5), indicating that polygenic risk contributes independently of APOE genotype [28].

### Neuroinflammatory and proteomic markers of resistance

Two cytokine principal components nominally discriminated resistant from high-pathology counterparts (Method 13): PC1 (*β* = 0.127, p = 0.046) and PC2 (*β* = 0.173, p = 0.052; Supplementary Table ST7; Supplementary Fig. S11; aligned in direction with [29]), though neither reached significance by ANOVA-based chi-square test (Fig. 3b). At the proteomic level (Method 14), alpha 1-antitrypsin (SERPINA1; P01009, p = 0.037) and fibrinogen alpha chain (FGA; P02671, p = 0.037) were both upregulated in resistant individuals (Fig. 3c; Supplementary Table ST4). This is consistent with an independent amyloid-PET-based proteomic analysis of the same cohort identifying both proteins, with SERPINA1 similarly elevated in amyloid-negative individuals [30]. SERPINA1 is a serine protease inhibitor implicated in neuroinflammatory regulation [31], while FGA upregulation has been linked to vascular and coagulation dysregulation in AD [32]. These proteomic and cytokine signals require replication in an independent cohort before definitive interpretation.

### Neuroinflammatory and genomic patterns of resilience

No PRS term distinguished resilient individuals from high-pathology MCI counterparts (all p *≥* 0.09; Method 12), nor did any cytokine PC after accounting for age and sex (all p *≥* 0.14; Supplementary Tables ST5–ST7; Method 13). Resilient individuals were, however, distinguished from high-pathology AD by APOE carriership (*β* = 0.48, p = 0.002; Supplementary Table ST5) and by all three cytokine components (PC1: *β* = 0.183, p = 0.004; PC2: *β* = 0.253, p = 0.004; PC3: *−*0.233, p = 0.042; Supplementary Table ST7; Supplementary Fig. S11). AD was enriched for *ε*4-containing genotypes, while resilient individuals showed greater representation of neutral and protective genotypes (Fig. 3a; *ε*3/*ε*4 reduction in resilience was nominally significant before multiple comparisons correction). This suggests that *ε*4 enrichment specifically marks the transition to clinically manifest AD, absent in the resilient-versus-MCI comparison. The positive PC1/PC2 associations indicate lower overall pro-inflammatory and chemokine burden in resilient relative to AD individuals; the negative PC3 association, dominated IL-20 and MIP-1/CCL3 loadings (Supplementary Fig. S11), suggests reduced tissue remodelling signalling alongside preserved monocyte chemotactic activity [33, 34]. No proteomic differences reached significance in either resilient comparison (Method 14), warranting replication in larger cohorts.

### Blood-based and A*β* PET-based pathological stratification

Blood-based biomarkers offer a scalable and minimally invasive readout of AD-related pathology, but capture peripheral rather than cortical protein dynamics. To test whether the BBM-defined resistant and resilient phenotypes are recoverable using cortical amyloid burden directly, and to characterize the relationship between peripheral and cortical pathology, we repeated the stratification profiling analysis by substituting A*β* PET positivity as the sole pathology axis (Fig. 4a; Extended Fig. 2; Methods 9–10).

**Fig. 4:**
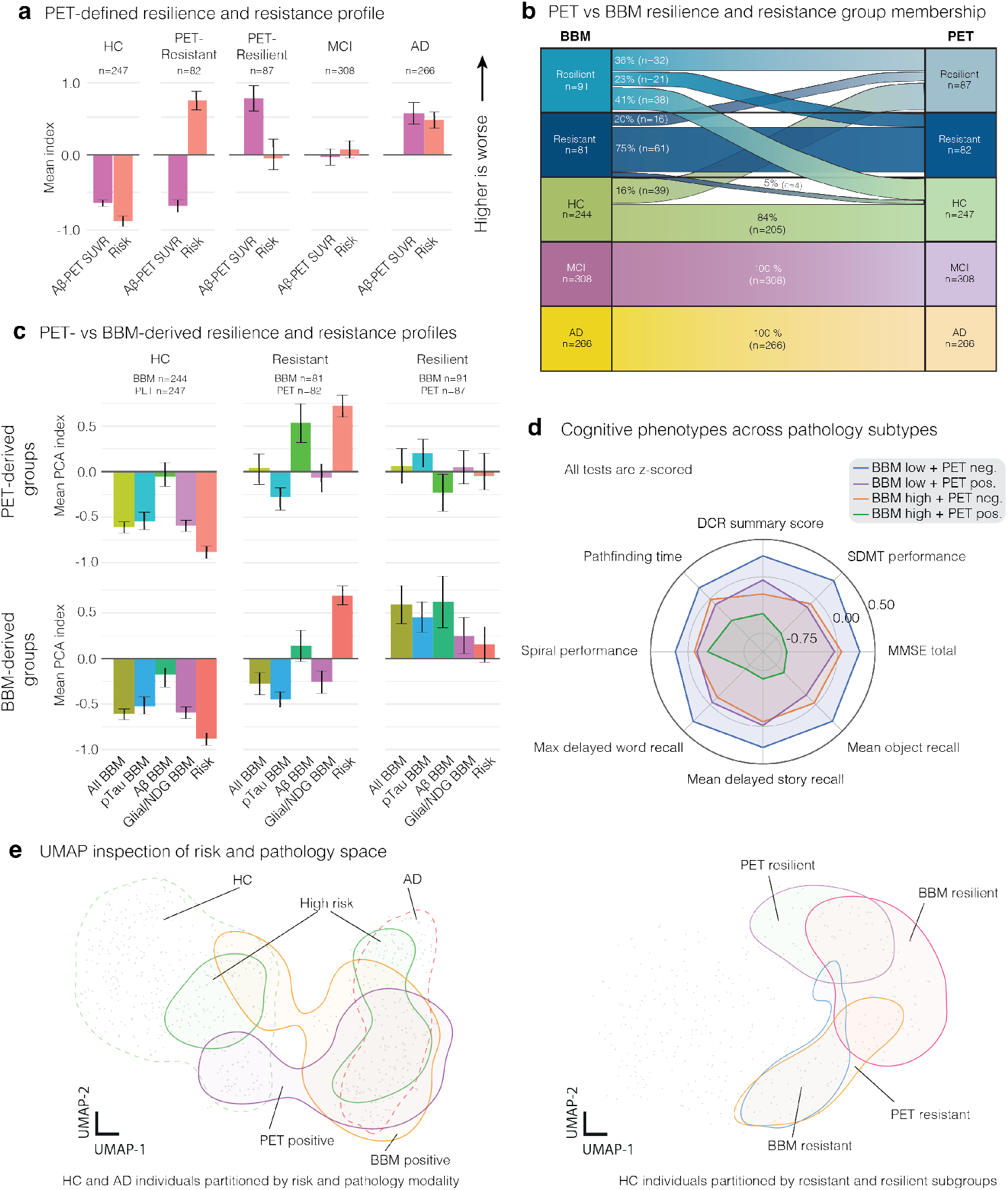
Neuropathological profiles of resilience and resistance defined by blood-based biomarker and PET modalities. a, PET-derived resilience/resistance profiles across HC, resistant, and resilient groups, comparing mean scores *±* s.d. (A*β*-PET SUVR and risk PCA index). As expected, resilient features high SUVR, while resistant features low SUVR and high risk. **b,** Sankey diagram of BBM-vs. PET-derived group correspondence. HC shows 84% concordance (n = 205), and resilient and resistant groups exhibit greater inter-modal discordance (36% and 75% concordance, respectively); MCI and AD classifications are fully concordant by construction. **c,** PET-and BBM-derived resilience/resistance profiles across HC, resistant, and resilient groups. Domain-specific mean scores *±* s.d. (All BBM, A*β* BBM, p-tau BBM, Glial/NDG BBM, and risk), for groups derived independently from PET (*top*) and BBM (*bottom*). Group sizes shown above each column. distinct domain-level differences between modalities, particularly for A*β* BBM, p-tau BBM, and risk axes. NDG = neurodegenerative. **d,** Cognitive performance (eight neuropsychological measures) across four BBM*×*PET concordance subgroups. BBM-high/PET-negative and BBM-low/PET-positive exhibit markedly similar cognitive profiles across the eight measures; BBM-high/PET-positive shows the most cognitive impairment. **e,** UMAP combined risk and pathology features (Method 11). *Left*, density contours for five groups across HC and AD individuals: Healthy (green, dashed), High Risk (green), AD (red, dashed), PET-positive (purple), BBM-positive (orange), showing partial overlap between diagnostic categories. *Right*, contours for BBM-resilient (pink), PET-resilient (purple), BBM-resistant (blue), and PET-resistant (yellow) subgroups, showing partially overlapping but distinct regions in HC individuals. MCI excluded from both UMAPs to improve visual distinction. Note that resistance and resilience are mutually exclusive; apparent contour overlap reflects proximity in the projected space, not shared membership.

### Modality-dependent differences in subgroup membership and profiles

Switching from BBM to PET altered subgroup membership systematically (Fig. 4b). Individuals with low blood pathology, including HC and BBM-Resistant participants, largely retained their assignments. BBM-defined resilience showed more substantial reclassification: 64% of BBM-Resilient individuals were reassigned to either PET-Resistant (23%) or HC (41%), reflecting elevated blood pathology alongside low or absent cortical A*β* burden (Fig. 4b). Conversely, approximately 20% of BBM-Resistant individuals were reclassified as PET-Resilient, reflecting low peripheral pathology alongside pathological cortical A*β* accumulation, consistent with impaired clearance from brain to periphery [27, 35]. Risk scores did not differ significantly between BBM- and PET-derived definitions of either phenotype (resistant: p = 0.684; resilient: p = 0.342; Fig 4c), indicating equivalent risk profiles despite distinct pathological compartments. The discordant subgroups (BBM-High/PET-negative and BBM-Low/PET-positive) occupied distinct regions of the of the pathology-risk space (Fig. 4e, *right panel*; Extended Data Fig. 2c) yet showed strikingly similar cognitive performance and diagnostic composition (Fig 4d; Extended Data Fig. 2a). This indicates that modality-specific discordance does not translate into clinically distinguishable cognitive phenotypes at the current time point.

### BBM and PET as partially orthogonal pathology axes. Agreement between

BBM- and PET-derived pathology classifications was modest (Extended Data Fig. 2b; ARI = 0.137, Supplementary Fig. S10d), consistent with heterogeneous correlations between PET SUVR and individual BBM measures: p-tau217 and p-tau181 showed the strongest positive associations (r = 0.51–0.65), while blood A*β* was negatively correlated with SUVR across vendors (r = *−*0.10 to *−*0.45; Supplementary Fig. S10b). This discordance may partly reflect the known limitations of blood A*β* relative to plasma p-tau species [15, 26, 27, 36], the peripheral sink hypothesis [27, 35], or with assay-specific A*β*42/40 effects and cross-vendor pre-analytical variability [37, 38]. This is unlikely to be the full explanation, however: adding PET SUVR to the BBM clustering solution reduced mean bootstrapped ARI from 0.63 (s.d = 0.11) to 0.53 (s.d. = 0.22) and rendered the bootstrap distribution bimodal (Supplementary Fig. S10c), indicating that cortical amyloid introduces a competing cluster structure rather than simply augmenting the BBM solution. BBM and PET therefore likely capture overlapping but partially distinct aspects of AD-related pathology, and A*β*-PET positivity should not be treated as the gold standard against which our BBM clustering is validated.

### Convergent identification of resistant and resilient subgroups

Despite this partial orthogonality, A*β*-PET positivity alone was sufficient to identify resistant and resilient subgroups within the canonical HC group (Fig. 4a–c). BBM-and PET-derived classification showed equivalent performance in distinguishing clinical groups: BBM achieved a sensitivity/specificity of 0.59/0.78 for AD versus HC, compared to 0.61/0.78 for PET, with no significant difference in error pattern across all three clinical comparisons (McNemar’s test: AD vs HC, p = 0.709; MCI vs HC, p = 0.891; AD+MCI vs HC, p = 0.677; Supplementary Fig. S10e). The UMAP projection of the combined pathology, risk, and cognitive feature space illustrates this partial overlap: PET-positive and BBM-positive regions are adjacent but non-identical (Fig. 4e, *left panel*).

Together, these findings establish that resistant and resilient phenotypes are identifiable across pathology modalities (ie. PET and BBM) with equivalent clinical discriminability, supporting their validity as biologically meaningful entities. The systematic discordance between peripheral and cortical pathology is informative. The modality of interrogation therefore shapes both subgroup composition and the specific pathological features that define resistance and resilience.

### A minimum blood biomarker and risk battery for resilience and resistance classification

#### Minimum blood biomarkers

Four blood biomarkers (A*β*40, p-tau217, p-tau181, and NfL) were sufficient to closely replicate the full-panel BBM pathology clustering (ARI = 0.56 *±* 0.04; Fig. 5a). Sequential backward elimination (Method 15) showed clustering agreement stabilized at four features, with additional markers yielding only marginal ARI gains at increased operational burden (features 5–11 each contributed less than *<*0.02 ARI on average while adding assay requirements; Fig. 5a; Extended Data Fig. 4a). A three-marker battery omitting A*β*40 (p-tau217, p-tau181, NfL) provided a further-reduced option with comparable agreement (ARI = 0.54). Both panels were stable across vendor combinations, with 5-fold cross-validated balanced accuracy of 82–84% across all 24 four-marker and 8 three-marker configurations; within 3 percentage points of the full-panel ceiling (Extended Data Fig. 4b).

**Fig. 5:**
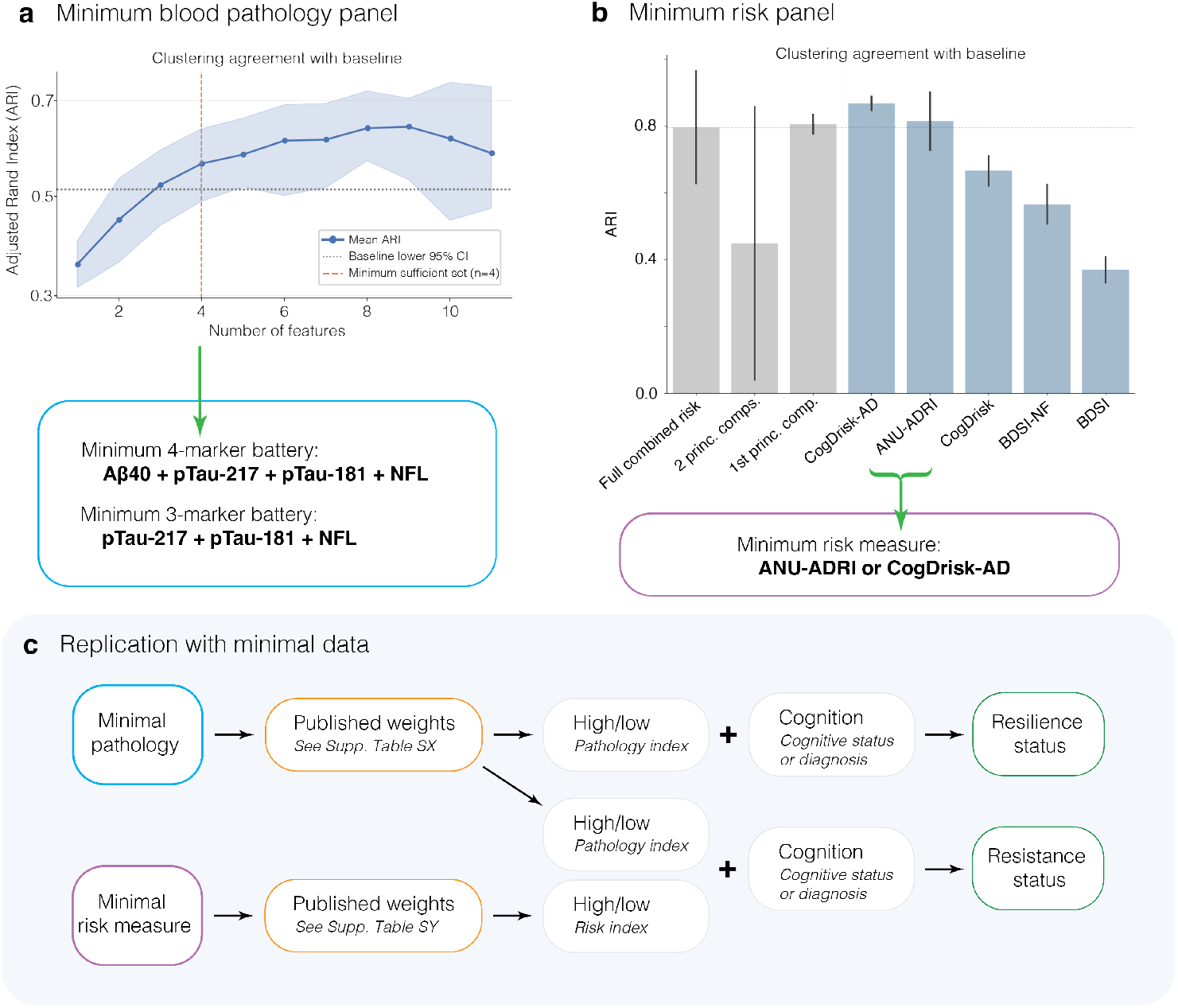
Minimal data requirements for replication of AD resilience and resistance classifications in the Bio-Hermes-001 cohort. a, Mean bootstrapped adjusted Rand index (ARI) [39] as a function of the number of blood biomarker features count, relative to a baseline (dotted line; Methods 7, 15). ARI stabilizes after four features (dashed vertical line), identifying a minimum BBM battery of A*β*40, ptau217, p-tau181, and NfL; a three-marker option (p-tau217, p-tau181, NfL) is also viable. Shaded band, 95% CI. **b,** Clustering agreement (mean ARI, *±* s.d.) with the full-panel solution across candidate risk measures (Method 16). The full combined risk score, CogDrisk-AD, and ANU-ADRI meet or exceed the baseline threshold (dotted line). **c,** Replication pipeline: minimal pathology data scored via published weights (Supplementary Table ST14) yield a high/low pathology index; minimal risk data scored similarly (Supplementary Table ST12; Method 17) to yield a high/low risk index. Combined with cognitive status or diagnosis, these enable resilience/resistance classification (Method 18).

#### Minimum risk scoring

CogDrisk-AD and ANU-ADRI each reproduced the full combined risk clustering at or above the baseline agreement threshold as single metrics (Fig. 5b; Extended Data Fig. 4c), making either a minimum sufficient risk measure for resistance classification (Method 16; Extended Data Fig. 4d).

#### Minimum replication modelling

To enable classification without re-running the full unsupervised pipeline, we provide two portable surrogate models (Method 17). For pathology, a logistic regression maps minimum-battery biomarker indices, each reconstructed from a single vendor-specific measurement via published centring parameters and PC1 loadings (Supplementary Table ST13), to the reference pathology label using Youden’s threshold (Supplementary Table. ST14). For risk, logistic regression on CogDrisk-AD or ANU-ADRI score reproduces the reference risk label (Supplementary Table ST12, ST11). Combining the resulting pathology and risk indices with cognitive status or clinical diagnosis recovers full classification at *≥*82% balanced accuracy (Fig. 5c). A three-or four-marker blood panel, a single validated risk instrument, and clinical cognitive assessment constitute a minimum replication toolkit for the phenotypes defined here (Method 18). Python and R code for our replication toolkit available at https://github.com/Dibz15/ad-resilience.

## Discussion

Applying a stratification framework across 19 plasma assays and six risk instruments in 990 participants, we identified resistant and resilient groups in the Bio-Hermes-001 dataset that are clinically and molecularly distinct from canonical HC, MCI and AD groups (Figs. 2, 4). Consistent with established APOE risk gradients [40, 41], the genomic profiles of resistance and resilience mapped onto the expected continuum of genetic protection: *ε*3/*ε*3 enrichment and attenuated polygenic risk in resistance, an intermediate profile in resilience, and peak *ε*4 burden in high-pathology AD [42, 43]. Resilient individuals lack this protective architecture but are similarly spared AD’s *ε*4 enrichment, distinguished from clinically impaired counterparts instead by a multicomponent neuroinflammatory cytokine profile (Fig. 3). This raises the possibility that clinical transition under high amyloid and tau burden depends on additional pathological processes, such that intervening on these processes, rather than amyloid and tau alone, could delay progression in individuals already carrying substantial core AD pathology. Distinct from resistant phenotypes, this gradient is potentially consistent with resilience representing a more proximal stage of AD-related biological progression [10, 44]. Whether these cytokines reflect an active neuroprotective mechanism or a bystander effect of pathological severity remains open for longitudinal investigation.

### Blood biomarkers and PET as distinct measures of resilience and resistance

Resistant and resilient phenotypes are not artefacts of a single pathology modality, remaining identifiable and equally discriminable using cortical A*β* PET positivity alone (Fig. 4; Extended Data Fig. 2). Agreement between blood-and PET-derived classification was nonetheless modest, likely reflecting the known limitations of blood A*β* relative to plasma p-tau. Notably, incorporating PET into a BBM clustering solution already dominated by p-tau introduced a competing cluster structure rather than reinforcing it (Supplementary Fig. S10c), a pattern broadly consistent with reports that plasma p-tau217/A*β*-PET weakens outside the extremes of the biomarker continuum [45–47]. Together, these findings suggest that peripheral and cortical measures of AD-related pathology capture partially distinct biological signals, neither offering a gold standard for validating the other. Methodological decisions for identifying clinical phenotypes should therefore account for the pathological signature of interest, treating cross-modal discrepancies as potentially informative rather than as noise.

### Operationalizing the identification of resilience and resistance

Amid growing interest in earlier prognosis [48], these findings demonstrate that resistant and resilient phenotypes, as operationally defined here, can be identified from a single cross-sectional blood biomarker assessment, offering a scalable alternative to definitions requiring years of longitudinal follow-up. Realizing this translational potential requires that the framework be operationalizable outside cohorts with the extensive multi-vendor coverage available here. We therefore identified a minimum sufficient battery (three to four blood-based markers and a single risk score) sufficient to recover the full classification (Fig. 5). This scalability supports two trial applications. First, screening for resilient individuals could allow exclusion or stratification of participants unlikely to decline despite high pathological burden from trials targeting amyloid or tau clearance in cognitively unimpaired populations, reducing dilution of treatment effect. Second, resistant individuals, who combine high estimated risk with low pathological burden and preserved cognition, represent a candidate population for primary prevention trials aimed at testing whether this low-pathology state can be sustained or extended.

### Grounding resilience concepts in multi-modal biomarker frameworks

Differences between resilient individuals and other clinical groups align with previous evidence of soluble A*β* accumulation in cortex and hippocampus [8] and elevated p-tau and TDP-43 proteotoxicity effectively proxied in blood [48, 49]. This sparing of cognition may reflect neuronal compensatory mechanisms, consistent with the distinct neuroinflammatory profile observed in resilient relative to high-pathology AD individuals. The more pronounced genomic differentiation of resistant individuals, evident even relative to high-risk clinical counterparts, contrasts with the more restricted genomic structure of resilience, present only in clinically manifest AD. This asymmetry is consistent with cognitive reserve being driven primarily by non-hereditary lifestyle factors rather than genetic protection [9, 50]. Grounding these categories in independent molecular and pathological measures also addresses a longstanding epistemological limitation of outcome-based definitions, whereby target constructs are defined by the very outcome through which they are recognised. Together, these results support a shift from univariate proxy operationalizations of symptomatology toward multi-modal, data-grounded representations of the mechanisms underlying early AD pathology.

### Current use of resilience and resistance constructs

At present, resilience and resistance remain primarily retrospective explanatory constructs in research, commonly operationalized via cognitive-reserve proxies such as education and occupational complexity, rather than as prospectively defined, biomarker-based categories applied at the point of participant selection [10, 11, 21]. Existing clinical trial enrichment strategies target the opposite end of the risk spectrum, identifying individuals most likely to decline: plasma p-tau217 and p-tau231 capture early amyloid pathology and have been proposed as enrichment markers for preclinical AD trials [51], and combined plasma p-tau217 with digital cognitive assessment has been used to identify individuals at elevated risk of near-term decline [52]. No existing framework prospectively identifies resilient or resistant individuals using molecular biomarkers for trial inclusion or exclusion, a gap this framework addresses.

### Limitations

The cross-sectional design cannot distinguish durable resistance or resilience from a transient state preceding rapid cognitive decline, nor establish the temporal sequence in which molecular markers, such as the cytokine profile distinguishing resilient from AD individuals, emerge relative to pathological accumulation. The derived resistant (n = 81) and resilient (n = 91) groups also remain modest in size for subgroup-level-omics analysis. A*β* PET was the only brain-derived measure available, preventing an equivalent cross-modal comparison for tau pathology. This is distinct from the observed blood-PET discordance discussed above. Cognition was defined by Bio-Hermes-001 investigator judgment rather than a performance composite matched in construction to our pathology and risk clustered composites, and functional independence was embedded in that judgment rather than modelled explicitly. None of these limitations should be difficult to address with an appropriately designed cohort.

### Future work

Future work should validate these groupings longitudinally to confirm disease trajectories align with theoretical expectations, incorporating both A*β* and tau brain-derived measures, ideally continuous rather than binary (e.g. centiloid), and confirm that the minimum sufficient battery identified here generalizes to independent cohorts. Expanded-omics profiling following this validation would support digital phenotyping and, ultimately, interventions to sustain resistance or enhance resilience.

Together, these findings extend prior cross-sectional identification of resistance and resilience, previously reliant on single-analyte blood markers, CSF, or PET, to a multivariate, blood-based framework that is both more comprehensive and deployable. The genomic gradient separating resistant, resilient, and clinically impaired individuals offers a concrete hypothesis for the biological machinery underlying protection against AD. Most immediately, the minimum biomarker and risk battery identified here, recovering the full classification at 82–84% balanced accuracy across vendor combinations, lowers the barrier to replicating and deploying this framework, from vendor-specific research assays to a panel usable in prospective cohorts and, eventually, clinical practice. Closing the diagnostic gap between molecular pathology and clinical presentation will require exactly this kind of translation: definitions of risk and protection that are biologically grounded, operationally minimal, and available before cognitive decline has occurred.

## Methods

P-values for each modality of analysis are reported following Bonferroni corrections where there are multiple comparisons with the same outcome and modality. Values are reported precisely, except where *p <.*001 where this notation is used instead.

### Method 1: Study aims and design

With the present study, we aimed to investigate the following exploratory objectives. Aim 1 was preregistered [53], while aim 2 and 3 were post hoc and not preregistered:

- Aim 1: Can resistant and resilient subjects be identified in this cross-sectional cohort using a theory-driven and data-grounded approach?
- Aim 2: Are the groupings meaningful as intended?
- Aim 3: What is the minimum battery of pathology measures required to replicate the stratification framework in other settings?

To identify the resistant and resilient groups, we relied on theoretical conclusions in published precedent. Accounting for the clinical classification of the cohort by the original study investigators (Supplementary Table ST3), pathology and risk were the last remaining factors required to identify the groups in this sample. As stated in our preregistration, we created a pathology composite and risk composite to find datadriven thresholds for the two variables required for stratification. In conjunction with the criteria represented in Table X, the cluster thresholds were used to distinguish resistant and resilient subjects in the clinically-defined cognitively healthy cohort.

To avoid circularity in the use of features for deriving the groups to validating the methodology, we selected the second most accessible biomarker [17] available in the dataset, that is AB PET, to explore whether the pathology composite based on blood biomarkers captures additional pathology types as would be theoretically anticipated. We compared the assignment of subjects to resistant and resilient groups to a random assignment to the same number of groups overall, to explore whether the composite approach created a representation theoretically more meaningful and practically distant from unbiased randomness.

### Method 2: Dataset

All our analyses were conducted using the Bio-Hermes-001 cohort [20], made available to us and other UK-based research teams via the Data Challenge [54] and accessed via the AD Workbench [55]. The cross-sectional cohort includes data from 1,009 older adults aged between 60–85 (56% female), recruited across 17 sites in the United States, with a priority objective to include individuals from populations typically underrepresented in dementia research. Some participants were excluded from our analysis (Method 4). Ethical approval for this secondary analysis was provided by the Data Challenge [54] via the University of Glasgow (application number: 200230198). Governance was in accordance with policies of the data guarantor, the Global Alzheimer’s Platform Foundation. We utilized assay results from Aural Analytics, C2N, Cognivue, IXICO, Lilly, Linus, Merck, Quanterix, Roche Diagnostics, University of Gothenburg, as well as results collected and analysed by the Bio-Hermes-001 study sites (Supplementary Table ST8). Units were converted as necessary for cohesion and all test results were standardized to the overall sample distribution for most analyses (Supplementary Table ST18).

### Method 3: Ethics statement

Written informed consent in compliance with relevant guidelines was provided by participants as part of the original Bio-Hermes-001 study [20]. The present secondary analysis of the dataset was approved as part of the University of Glasgow-led Data Challenge (College of Medical Veterinary and Life Sciences Ethics Committee, application number 200230198).

### Method 4: Exclusions and missingness

From the original cohort, only individuals with an adjudicated cognitive classification (healthy cognition, mild cognitive impairment or probable AD) according to the Bio-Hermes-001 criteria (n = 1,001 of the full 1,009), were included in our clustering analyses. Individuals with incomplete data for the variables included in our composite measures of pathology were excluded (final n = 990). For the pathology composite, where missingness was due to a value being out of range of detection, we imputed a value accordingly: where below the threshold we imputed the minimum and where over the threshold the maximum value observed in the sample overall. Additionally, some biomarkers were winsorized to account for extreme outliers (Supplementary Table ST9). Missingness was retained in values incorporated into the risk composite (Methods 5, 6b). We excluded cytokines with over 70% missingness and proteins with over 50% missingness (Methods 13, 14). The protein panel abundance threshold is more conservative because these protein blood panels have greater precedent of use in research in comparison to cytokine measures.

### Method 5: Risk measures

Risk measures, as described in this work, are instruments from the literature for stratification of individual risk for the development of dementia. These instruments use demographic, and lifestyle factors such as age, sex, education to determine a single “risk score” for the individual, though the included features and the final range of outcomes depends significantly on the specific instrument [12, 13]. As described in [56], we reviewed literature on dementia-related risk score calculations, and we implemented six risk scores from the literature: the Cardiovascular Risk Factors, Aging, and the Incidence of dementia (CAIDE; [57]) with the previously validated APOE-e4 alle carriership adaptation [58]; Lifestyle for BRAin Health (LIBRA; [59, 60]) with Geriatric Depression Scale-measured depression [61, 62]; Brief Dementia Screening Indicator (BDSI; [63]); Australian National University AD Risk Index (ANU-ADRI; [13, 64]), Cognitive Heath and Dementia Risk Index (CogDrisk) and CogDrisk for Alzheimer’s Disease (CogDrisk-AD; [13]). The complete list of risk scores assessed, adopted, and detailed justification for method of implementation in Bio-Hermes-001 are previously presented [56]. Note that in some of our analyses, where listed, a seventh modified risk score was used; BDSI-NF, which is the aforementioned BDSI index without the functional component.

### Method 6: Composite variables via unsupervised clustering

#### a. General approach

To accomplish aim 1, it was determined that we would need a label-free (unsupervised) approach for identifying “high” and “low” BBM pathology and risk levels. Risk and pathology were each analysed separately, though they each used a similar clustering approach to produce the cluster labels. To prepare the data for analysis, we standardized all non-categorical variables. We conducted clustering with both k-means [65] and Bayesian Gaussian Mixture Models (BGMM; [66]), and qualitatively compared their clustering along the underlying features. For both pathology and risk, BGMM was the most suitable for the a priori planned two-level classification (low-high). This approach was selected for the group assignment over three or more levels of the composite measures, to avoid further complication in defining thresholds in the absence of related precedent. Therefore, we used BGMM 2-level (high-low) classification to assign into the final groups in order of theoretical progression and according to the predetermined theoretical definition (HC/Resistant/Resilient/MCI/AD). Silhouette scores confirmed that this provided the most distinct clustering in each case (Extended Data Fig. 5).

Permutation testing confirmed that our clustering separation exceeded chance (Supplementary Tables ST1, ST2). Several biomarkers exhibited non-linear divergence near the cluster boundary rather than a simple threshold, supporting multivariate clustering over univariate cut-points (Supplementary Figs. S1–S7). Clustering used only blood-based pathology and demographic risk measures; cognitive and diagnostic status was determined independently by clinical adjudication.

#### b. Risk composite

We created the risk composite using the scores calculated according to seven indices, each standardized to its distribution. The risk indices captured age, sex, years in education, hypertension and other risk factors according to validated algorithms operationalized to be applied in the Bio-Hermes-001 dataset (Method 5). For the BGMM algorithm, “tied” covariance was selected using the lowest BIC across covariance settings (Extended Data Fig. 5b).

#### c. Pathology composite

We created the pathology composite using the standardized values of the AD pathology blood biomarkers with complete results following imputation of out of range values (Method 4). The pathology composite incorporated 19 features, including amyloid beta, phosphorylated tau, and other markers from different vendors (Supplementary Table ST8). For the BGMM algorithm, “full” covariance was selected using the lowest BIC across covariance settings (Extended Data Fig. 5a).

#### d. Null clustering permutation testing

To confirm that the resistant and resilient profiles reflect structured biological separation rather than an artefact of the clustering procedure, group assignments were compared against random permutations of the pathology and risk cluster labels; the observed profile separation significantly exceeded the permutation-derived null distribution in both cases (Supplementary Tables ST1, ST2).

### Method 7: Clustering similarity measurement

Comparison of BGMM-derived clusterings across different feature sets and conditions (Method 6) required a principled measure of both inter-clustering similarity and within-clustering stability. Pairwise similarity between distinct clusterings was quantified using the adjusted Rand index (ARI) [39], defined on (*−*1, 1], where 1 denotes perfect agreement and 0 denotes agreement at chance level. Negative values, indicating worse-than-chance agreement, are rare in practise. Importantly, because the ARI is adjusted for chance under the hypergeometric distribution, it accounts for label imbalance; in our dataset, where the positive cluster comprises approximately 28% of the sample, a näıve overlap metric would be inflated by majority-class agreement, whereas the ARI remains interpretable across cluster size asymmetries. As a reference, an ARI of *∼*0.5 in a dataset with this imbalance reflects substantially better-than-chance agreement and not merely majority-class overlap.

To assess clustering stability, we employed a bootstrapped similarity procedure. For a given clustering *C* derived from the full sample, we generated *B* bootstrap resamples, re-fitted the clustering model on each, and computed the ARI between each bootstrap clustering and *C* as a static reference. Stability was then summarised by the bootstrap mean and standard deviation:

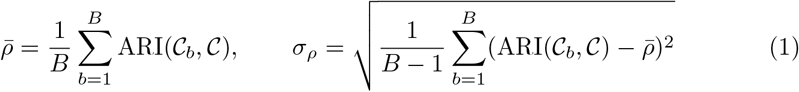

where *C_b_* denotes the clustering obtained from the *b*-th bootstrap resample and *B* is the number of iterations specified in the corresponding method. This procedure was applied both to quantify self-similarity of composite clusterings and to compare clustering agreement across reduced feature sets.

### Method 8: Construction of PCA-derived indices

#### a. Derivation

In some cases where it was needed to compare against large groups of features, we performed Principal Component Analysis (PCA; [66]) to create a proxy index of those features using the first principal component. To encourage uniform scaling when comparing multiple of these PCA-derived indices, we divided each first component by the square root of its eigenvalue to update its standard deviation to 1 (standardized PCA).

#### b. Comparison indices

We utilized these PCA indices to generate a unified axis for all pathology biomarkers in the pathology composite and separately for A*β*, p-tau, and other markers in the composite measure, as well as all indices in the risk composite; and numerous cognitive performance markers that were not involved in the clinical definition of the original study subgroups (see Supplementary Table ST15, Supplementary Fig. S8 for a full description of each PCA index). These axes were used for descriptive and inferential comparison of the final five groups, including measures of central tendency and spread with standard deviation bars (Figs. 2, 4).

#### c. Minimum blood biomarker battery selection indices

In the case of our minimum biomarker battery selection pipeline, it was desired to represent each distinct blood biomarker by only one feature, rather than each individual vendor-specific feature. As such, we created a PCA-derived index feature for A*β*40, A*β*42, p-tau181, p-tau217, NfL, and GFAP. In each case, the first component represented at least 60% of the group’s variance. These indices were then used in the subsequent minimum battery analysis and selection pipeline. See Supplementary Table ST13 for a description of each index for this analysis.

### Method 9: Blood-and PET-based pathology comparison

To characterize the relationship between A*β*-PET and BBM composite positivity (Method 6c), we examined their agreement with each other and their concurrent validity against cognitive and diagnostic status.

#### a. A*β*-PET measures

In the Bio-Hermes-001 dataset, A*β*-PET data were available from both IXICO and Eli Lilly protocols. IXICO data were selected because both a binary clinical label (A*β*-PET positive/negative) and a continuous standardized uptake value ratio (SUVR) were available. IXICO SUVR and Lilly centiloid values were found to be highly linearly correlated (Supplementary Fig. S10a), supporting their interchangeability for the purposes of this comparison.

### b. A*β*-PET and BBM biomarker relationships

The association between A*β*-PET SUVR and the individual blood biomarkers comprising the BBM composite was assessed using two complementary approaches. First, Spearman rank correlations between SUVR and each vendor-specific biomarker feature were computed directly (Supplementary Fig. S10b). Second, participants were stratified into four groups defined by the cross-classification of BBM and PET positivity status, and within-biomarker *z*-scores were compared across groups for all 19 biomarker features (Extended Data Fig. 4e).

#### c. Combined BBM and A*β*-PET clustering stability

Bootstrapped clustering stability (Method 7) was assessed for both the full BBM-only composite (Method 6) and a distinct composite derived from BBM features combined with A*β*-PET SUVR, to evaluate the marginal contribution of PET to clustering structure (Supplementary Fig. S10c)).

#### d. Concordance with clinical diagnosis and cognitive status

The distribution of clinical diagnosis labels was compared between participants stratified by BBM composite status and those stratified by A*β*-PET clinical label. Sensitivity and specificity for the prediction of diagnostic label (e.g., AD versus cognitively healthy) were computed for both stratification methods and compared. Agreement between BBM-and PET-defined pathological status was assessed using McNemar’s test, which evaluates whether the two methods produce statistically equivalent patterns of discordance (Supplementary Fig. S10e). Mean performance across eight cognitive tests was compared across the four cross-classified BBM-by-PET groups (Extended Data Fig. 4c).

### Method 10: Blood-and PET-based group comparison

Since normality of the standardized pathology axis scores was not assumed a priori, the Shapiro Wilk test was used to assess normality within each group, and Levene’s test was used to assess homogeneity of variance between groups. Where both assumptions were satisfied, group means were compared using a Student’s t test, or Welch’s t test when variances were unequal. Where normality was violated, a Wilcoxon rank sum test was used instead. Effect sizes were quantified using Cohen’s d, taken as an absolute value, with the direction of each effect recorded separately according to the sign of the difference between group means. Results are visualized in Fig. 4a, Extended Fig. 2, and in Supplementary Table ST16.

### Method 11: Risk-pathology space UMAP visualization

To visualize the distribution of clinically and biologically defined participant sub-groups within a shared biomarker feature space (Fig. 4e), we projected multi-modal biomarker data into two dimensions using Uniform Manifold Approximation and Projection (UMAP; [25]). The embedding was constructed from standardized principal-component proxies for plasma A*β*40, p-tau217, p-tau181 and NfL, together with amyloid PET SUVR (z-scored), a composite risk score (Methods 8, 5), and diagnostic status (AD-positive indicator), yielding a joint representation of amyloid, tau, neurodegeneration and clinical risk axes. Participants with a diagnosis of mild cognitive impairment were excluded from each projection to emphasize contrasts between cognitively healthy and clinically affected (AD) endpoints. For each binary subgroup of interest (e.g., cognitively resilient, cognitively resistant, PET-defined resilient/resistant, amyloid-, tau-, and risk-cluster-positive individuals), we estimated a two-dimensional kernel density (Gaussian KDE, Scott’s rule) over the shared UMAP coordinates using only members of that subgroup, and overlaid a single contour reflecting the region containing the highest-density mass of positive cases. Because empty regions of the embedding grid otherwise produce spuriously permissive density percentiles, contour thresholds were computed relative to the density evaluated at each subgroup’s own points rather than across the full grid, with the percentile threshold tuned per subgroup (15–40th percentile) to account for differences in subgroup prevalence and dispersion. All background participants were plotted as a low-opacity scatter to preserve visual context of the full cohort. Note that resistant and resilient groups are mutually exclusive by definition; apparent contour overlap reflects proximity in the projected space, not shared membership.

### Method 12: Polygenic risk scores

For genomics, we performed linear regressions and obtained Area Under the Curve (AUCs) for PRSs and APOE to explore the discriminatory power of each genetic profile marker. To assess genomic differences, we created Polygenic Risk Scores (PRSs) with Continuous Shrinkage (PRS-CS) only for subjects from genetically only European and near European populations (n = 770; Method 4). We adjusted PRSs for five PCs to represent population structure, and performed four genome-wide associations (GWAS) for: clinical AD with and without the APOE region of chromosome 19:44-46.5Mb) [67]; proxy AD [68]; educational attainment [69]; and cognition [70]. We represented APOE by the number of *ε*2 and *ε*4 alleles, as in [67]. APOE and all PRSs were adjusted for the population PCs before standardization.

### Method 13: Cytokine analyses

For neuroinflammation, we performed linear regressions for each outcome using principal components to reduce dimensionality and investigate model performance in explaining variance. We created three PCs to reduce dimensionality of the abundant cytokine panel results, which were rotated using varimax to maximize interpretability, following the measurement and statistical methods from [29]. The first PC reflecting general neuroinflammation is predominantly determined by upregulated cytokines IL16, eotaxin CCL11, TNF RII, IP10 CXCL10. The second PC reflects proinflammatory activation, inncluding upregulated ENA 78 CXCL5, IL17A CTLA B, IL 7, and downregulated TNF RII, IP10 CXCL10, IL16. Cytokine abundance, as measured by the three PCs, were used as predictors in logistic regression models to distinguish between our three primary comparison groups. We adjusted cytokine abundance levels for age and sex (Supplementary Table ST7; Supplementary Fig. S11).

### Method 14: Proteomic analyses

We did not reduce dimensionality across 294 abundant proteins, as it would not meaningfully represent their broad range of function. We adjusted protein abundance levels for age and sex before standardization. To identify upregulated or downregulated proteins, we performed t-tests on the same binary outcomes as used in genomic and cytokine linear regressions (Methods 12, 13) and Bonferroni corrected p-values for each outcome comparison.

### Method 15: Minimum blood biomarker battery selection pipeline

To identify the minimum set of blood biomarkers whose clustering approximates that derived from the full biomarker panel, we developed a three-step sequential selection pipeline grounded in bootstrapped clustering similarity (Method 7). The sufficiency criterion was that a reduced-feature clustering should achieve a bootstrapped mean ARI with respect to the full-feature BGMM clustering approaching the self-similarity of the full-feature clustering under resampling. As described in Method 8c, biomarkers with measurements from multiple vendors were first consolidated into a single index per biomarker type (A*β*40, p-tau-181, etc.) via the first principal component, yielding 11 features as input to the selection pipeline. Note that these six of these features (A*β*40, A*β*42, p-tau181, p-tau217, GFAP, NfL) are PC1 indices, each derived from multiple vendor-specific measurements as described in Method 8 and Method 17a. They are not raw biomarker values or simple z-scores of individual assay measurements.

The pipeline proceeded in three steps. **Step 1 — backward feature elimination.** Starting from the full set of 11 biomarker indices, we iteratively removed the feature whose removal least reduced *ρ̄*, evaluated over *B*_1_ = 50 bootstrap iterations, yielding a sequence of reduced panels. The resulting *ρ̄*-versus-panel-size curve (Fig. 5a) was used to identify the approximate minimum panel size that preserved full-panel clustering fidelity. Balancing this against the operational objective of minimising panel size, we selected four as the practical minimum. **Step 2 — exhaustive combinatorial search.** We enumerated all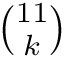combinations of *k ∈ {*3, 4, 5*}* indices and evaluated each by the scoring criterion

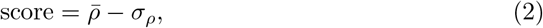

with *ρ̄* and *σ_ρ_*computed over *B*_1_ = 50 bootstrap iterations as defined in Method 7. **Step 3 — high-resolution re-evaluation.** The top 10 combinations by score were re-evaluated with *B*_2_ = 200 bootstrap iterations, and the combination with the highest score was selected as the final minimum battery (Fig. 5, Extended Data Fig. 4).

Note that the final selection of four as the practical minimum panel size reflects both the statistical stabilization of clustering agreement at this point and a deliberate operational judgment that the marginal gains in ARI from including additional markers do not justify the increased burden of a larger panel for future operationalization. We further provide a three-biomarker minimum panel where that is the more practical alternative.

### Method 16: Minimum operational risk score selection

To identify the minimum set of risk scores that adequately represents the full-panel risk clustering, we adopted an approach analogous to, but distinct from, that in Method 15. We first applied PCA to the full set of seven risk scores (Method 5); inspection of the component loadings (Extended Data Fig. 4d) revealed that the first two principal components captured the majority of total variance, with each component driven predominantly by a distinct subset of scores: PC1 was weighted primarily (in descending order) by COGDrisk, COGDrisk-AD, ANU-ADRI, BDSI-NF, and BDSI; and PC2 was weighted entirely by LIBRA and CAIDE. This bipartite loading structure motivated partitioning the seven scores into two groups aligned with PC1 and PC2, respectively. We then performed BGMM clustering independently on each group and evaluated the agreement of each reduced clustering with the full-panel clustering via bootstrapped mean ARI, using the same bootstrap procedure as in Method 15 (Fig. 5b).

A two-group partition was considered sufficient if the bootstrapped mean ARI of at least one group was comparable to the self-similarity of the full-panel clustering under resampling. The PC1 group met this criterion while the PC2 group did not, indicating that PC1-weighted scores dominate the variance structure of the original clustering and constitute its primary separating axis. To further assess interchangeability within the PC1 group, we additionally evaluated each constituent score individually (Fig. 5b).

### Method 17: Reproducible surrogate models for pathology and risk labels

To enable reproduction of our unsupervised pathology and risk labels in this and external cohorts without re-derivation of the full BGMM pipeline (Method 6), we provide a portable surrogate for each label. Throughout, the reported static BGMM clusterings (Method 6c) serve as the fixed reference labels; the models below are intended as faithful, reproducible reconstructions of those references, not as independent ground truth.

#### a. Pathology label via a surrogate logistic model

To assign the BBM composite pathology label without re-running BGMM clustering, we fit a logistic regression mapping the minimum-battery PCA indices (Method 15) to the static reference label. Each index is derived from vendor-specific measurements via a two-step transformation: each raw feature is first centred using its published per-feature mean, then multiplied by its published PC1 loading (Supplementary Table ST13), yielding a single index value per biomarker. Critically, the centring parameters and loadings are specific to each vendor-feature combination and are not interchangeable across vendors; replicating cohorts should apply the parameters corresponding to their assay platform, or use the recommended single-vendor proxy where only one platform is available. The logistic model is specified as

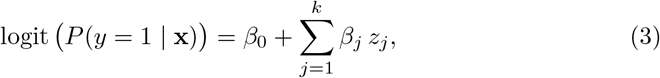

where *z_j_* is the standardized value of index *j*, *y* is the reference pathology label, and *k* is the number of distinct model features (ie. different biomarkers). Point estimates and model-based 95% confidence intervals are reported for each *β_j_* (Supplementary Table ST14). To recover balanced classification from the logistic output under the imbalanced positivity rate of the reference label, a Youden-index probability threshold (Supplementary Table ST14) is applied in place of the default 0.5 decision boundary. This threshold was selected to maximize balanced accuracy on the reference label and should be applied as reported for direct replication; cohort-specific recalibration may be appropriate where label distributions differ substantially from those reported here. Surrogate faithfulness was quantified by the cross-validated AUC for recovery of the reference label.

#### b. Risk label via single-score thresholds

Motivated by the close reproduction of the full risk clustering by individual risk scores (Method 16; Fig. 5b), we provide a logistic model of each individual risk score to predict the reference risk label (Method 6). These models were derived using the same method as for the pathology features (above), but using the risk features. Full parameters and balanced accuracy are reported in Supplementary Table ST12.

### Method 18: Operational definition of resilience and resistance

#### a. Theoretical definitions

Our operational definitions of each subgroup are derived from extensive literature on each and their corresponding theoretical and empirical characteristics [7, 10, 11, 21, 24]. As such, we refer as resilient to mean those who have a high (relative to other cognitively healthy individuals) level of measured pathology despite their cognitively healthy status, and resistant to mean those who have a low level of measured pathology and high level of estimated risk. In this study, we proxy *in vivo* pathology separately using two distinct primary modalities: composite blood plasma measures (Method 6) and A*β*-PET positive classification. Risk is proxied as a composite label via risk instruments from the literature (Method 5. Cognitive status is determined via clinical adjudication which was performed in the Bio-Hermes-001 cohort independent of our own analyses.

#### b. Operational Axis definitions

We define resilience and resistance operationally from the cross-classification of three independent axes: pathology, risk, and cognition. Each axis is assigned per participant using a reproducible procedure, such that the phenotypes can be reconstructed in this and external cohorts. The pathology and risk axes are assigned using the surrogate models of Method 17, which reproduce the corresponding BGMM reference labels (Method 6). For the pathology axis, the full transformation pipeline proceeds as follows: (i) centre each vendor-specific measurement using the published per-feature mean; (ii) multiply by the published PC1 loading to obtain the biomarker index (Supplementary Table ST13); (iii) stack the four index values and evaluate the logistic model 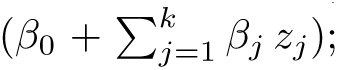 (iv) apply the published Youden-index probability threshold to obtain the binary high/low label (Supplementary Table ST14). For the risk axis, the pipeline is reduced to just applying the logistic model weights and published Youden-index probability threshold to the raw CogDrisk-AD or ANU-ADRI score (Supplementary Table ST12). The cognitive axis is left to the replicating investigator and may be operationalized using either clinical diagnostic status or a cognitive performance criterion, as available in the target cohort. In this study we used the clinical AD diagnosis label provided by the dataset creators.

#### c. Phenotype assignment

Each participant is assigned a binary status on each axis (using the aforementioned modelling), and resilience and resistance are yielded from the resulting cross-classification: resistant individuals are risk-positive but pathology-negative and cognitively healthy, and resilient individuals are pathology-positive but cognitively unimpaired.

### Method 19: Software

We used R via RStudio and Python in Jupyter as provided on the ADDI workbench with data access [55]. All machine learning models were implemented using scikit-learn [71].

## Data Availability

The Bio-Hermes-001 dataset (Clinical Trials ID: NCT04733989) is currently hosted on Alzheimers Disease Data Initiative (ADDI, report doi: 10.1002/alz.70278)), where it can be accessed upon request (https://discover.alzheimersdata.org/catalogue/datasets/8395cc96-f249-40db-beaa-4668feb3cc5e).

## Author contributions (CRediT)

**Kalliopi Mavromati**: Conceptualization, Data Curation, Formal Analysis, Funding Acquisition, Methodology, Project Administration, Resources, Software, Validation, Visualization, Writing – Original Draft, Writing – Review & Editing. **Connor Dalby**: Conceptualization, Data Curation, Formal Analysis, Methodology, Project Administration, Software, Validation, Visualization, Writing – Original Draft, Writing – Review & Editing. **Austin Dibble**: Conceptualization, Data Curation, Formal Analysis, Methodology, Project Administration, Software, Validation, Visualization, Writing – Original Draft, Writing – Review & Editing. **Ganna Leonenko**: Formal Analysis, Methodology, Software, Writing – Review & Editing. **Katherine Birditt**: Formal Analysis, Methodology, Software, Writing – Review & Editing. **Stelios Lamprou**: Formal Analysis, Methodology, Software, Writing – Review & Editing. **Lucie Tvrda**: Conceptualization, Methodology, Writing – Review & Editing. **Ioannis Konstantinidis**: Conceptualization, Methodology, Writing – Review & Editing. **Lynne Hughes**: Resources, Writing – Review & Editing. **Maura Malpetti**: Methodology, Supervision, Writing – Review & Editing. **Valentina Escott-Price**: Methodology, Supervision, Writing – Review & Editing. **Monika Harvey**: Methodology, Supervision, Writing – Review & Editing. **Alessio Fracasso**: Methodology, Supervision, Visualization, Writing – Review & Editing. **Michele Svanera**: Conceptualization, Methodology, Supervision, Visualization, Writing – Original Draft, Writing – Review & Editing. **Terrence Quinn**: Conceptualization, Methodology, Resources, Supervision, Visualization, Writing – Original Draft, Writing – Review & Editing.

## Code availability

https://github.com/dibz15/ad-resilience

## Extended Data

**Extended Data Fig. 1:**
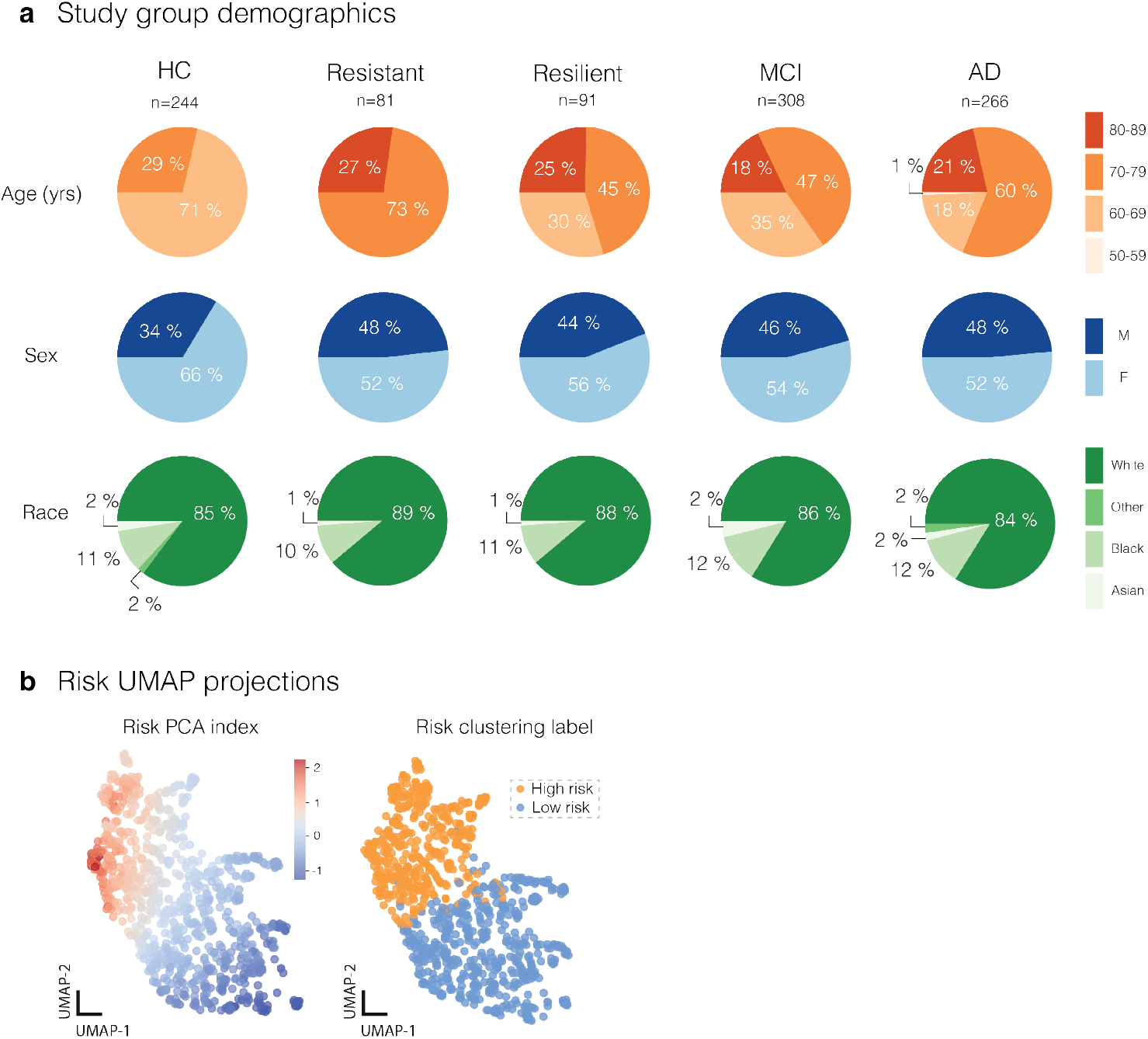
Study group demographics and risk clustering. Demographic and risk clustering profiles for five cognitive groups: healthy cognition (HC; n = 244), cognitively resistant (Resistant; n = 81), cognitively resilient (Resilient; n = 91), mild cognitive impairment (MCI; n = 308), and Alzheimer’s disease (AD; n = 266). a, Study group demographics. Pie charts show the distribution of age (50–59, 60–69, 70–79, and 80–89 years), sex (female, F; male, M), and self-reported race (Asian, Black, Other, White; categories as defined by Bio-Hermes-001) within each group. b, Risk UMAP projections. UMAP embeddings of demographic risk profiles across all individuals, colored by the continuous Risk PCA index (Method 8; left) and by a two-cluster solution distinguishing high-risk versus low-risk individuals (Method 6; right). The Risk PCA index is descriptive only and was not used to derive the clustering.

**Extended Data Fig. 2:**
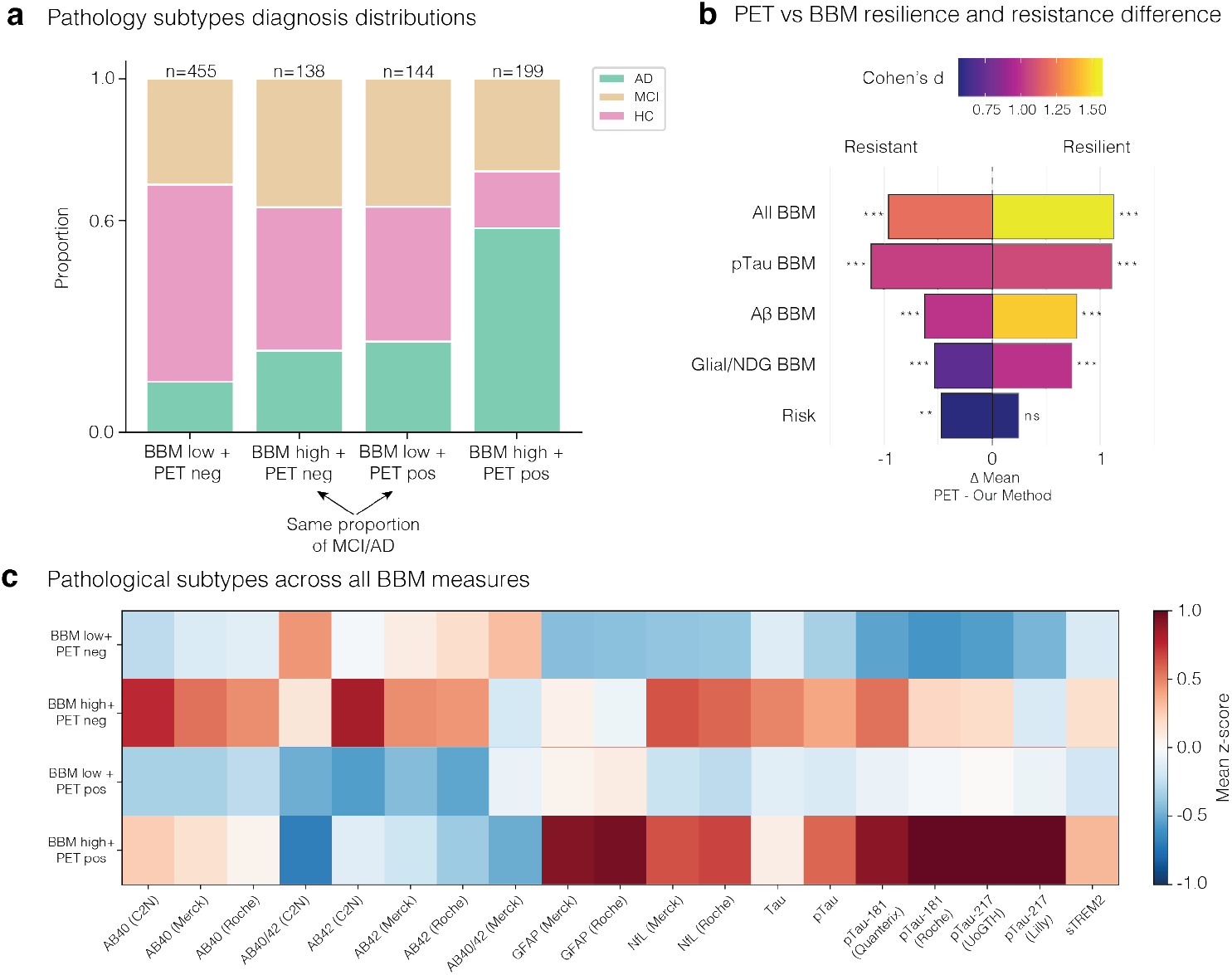
Cross-modal quantification of resilience and resistance discordance and characterization of pathological subtypes.a, Stacked bar charts showing the diagnostic composition (AD, green; MCI, tan; HC, pink) of each BBM × PET pathological subtype (n = 455, 138, 199, and 144, respectively), illustrating a similar distribution of diagnostic categories in the BBM high + PET neg and BBM low + PET pos subgroups, and the highest proportion of AD diagnoses in the BBM high + PET pos subgroup. b, Horizontal bar plots showing the mean difference in pathological domain scores (ΔPET − BBM method) for resistant (left) and resilient (right) individuals across five domains (All BBM, p-tau, Aβ, Glial/NDG, Risk), with bar colour encoding Cohen’s d effect size (scale 0.75–1.50). Significant differences are observed across all pathological domains (***p < 0.001; **p < 0.01), except for the Risk domain in resilient individuals (ns; Supplementary Table ST16). c, Heatmap of mean z-scores for a panel of blood-based biomarkers (Aβ40, Aβ42, GFAP, NfL, Tau, p-tau species, and sTREM2, measured across multiple vendors) across the four BBM × PET subtypes, revealing that the four subgroups have distinct blood pathology profiles.

**Extended Data Fig. 3:**
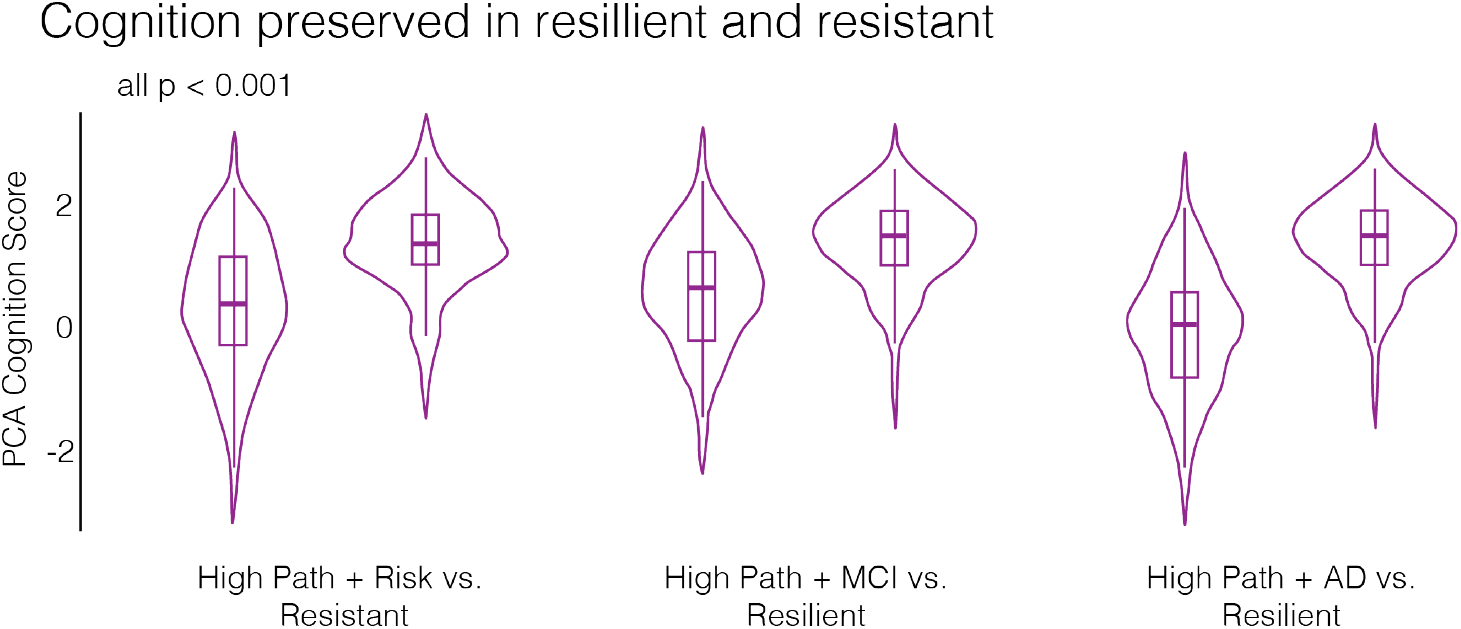
Verification of cognitive preservation in resilience and resistance. As expected by our counterfactual group definitions, cognitive performance was significantly higher in resistant and resilient individuals in all three comparisons (p < 0.001, two-sided Wilcoxon rank-sum test).

**Extended Data Fig. 4:**
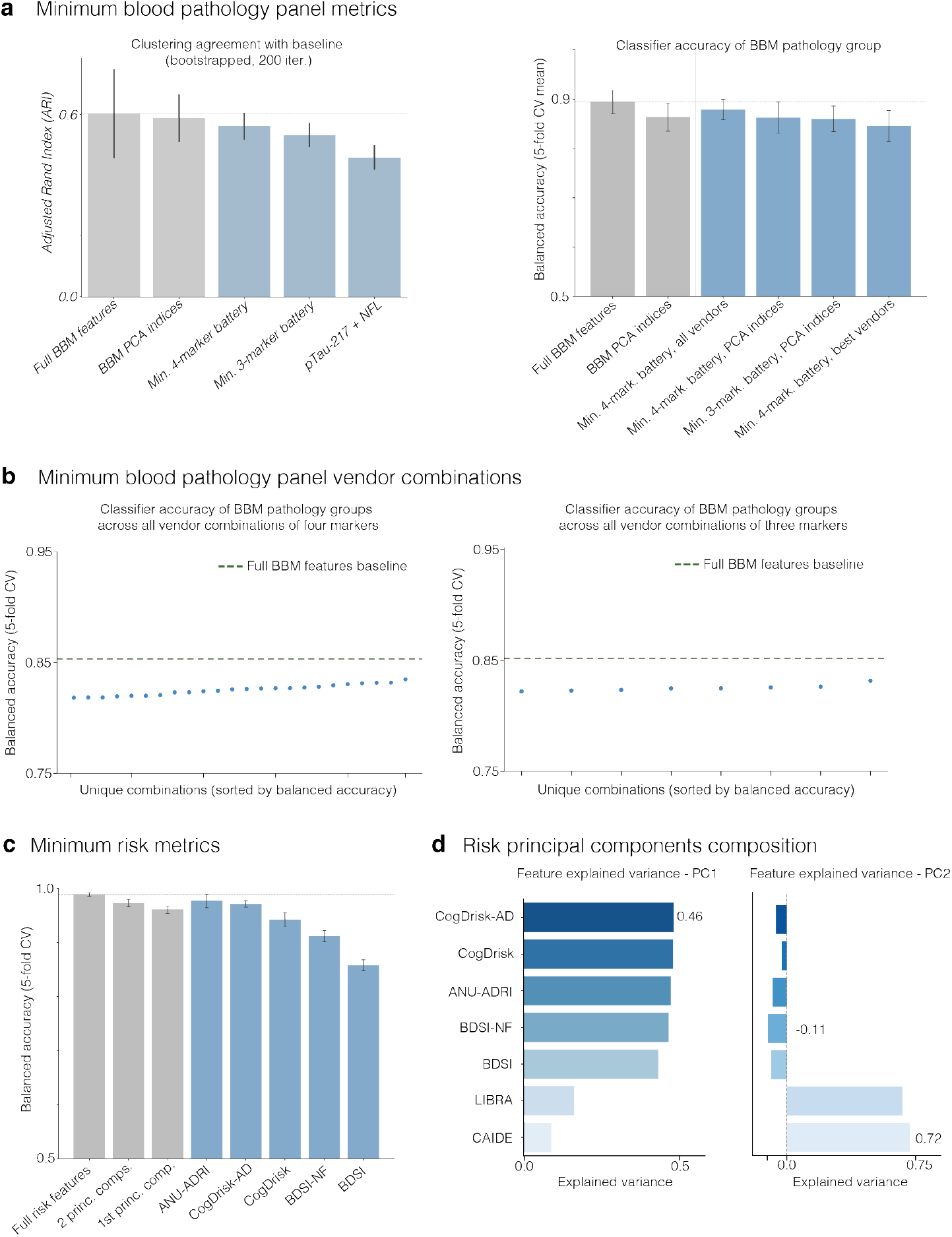
Minimum blood biomarker and risk metric panels for AD resilience and resistance classification.a, Left: clustering agreement (Adjusted Rand Index, ARI; mean ± s.d., 200 bootstrap iterations) between progressively reduced blood biomarker measure (BBM) panels —including the full BBM feature set, BBM PCA indices, minimum 4-marker battery, minimum 3-marker battery, and p-tau217 + NfL —and the full-panel baseline, corroborating the stability of the minimum sufficient batteries identified in Fig. 5. Right: balanced classification accuracy (5-fold CV mean ± s.d.) of BBM pathology groups derived from reduced panels, evaluated across all-vendor and best-vendor configurations, demonstrating that classification performance is largely prese3r7ved across panel reductions. b, Balanced classification accuracy (5-fold CV) across all unique vendor combinations for the 4-marker (left) and 3-marker (right) minimum BBM panels, sorted by performance; the dashed line indicates the full BBM features baseline, illustrating that most vendor combinations approach but do not reach full-panel accuracy, with a maximum delta of-3%. c, Balanced classification accuracy (5-fold CV, mean ± s.d.) for individual risk instruments and principal component-based reductions relative to the full risk feature set, validating that CogDrisk-AD and ANU-ADRI retain near-baseline performance as single-instrument minimum inputs. d, Loadings of the seven dementia risk scores on the first (PC1) and second (PC2) risk principal components. This identifies that there are two unique groups of risk scores.

**Extended Data Fig. 5:**
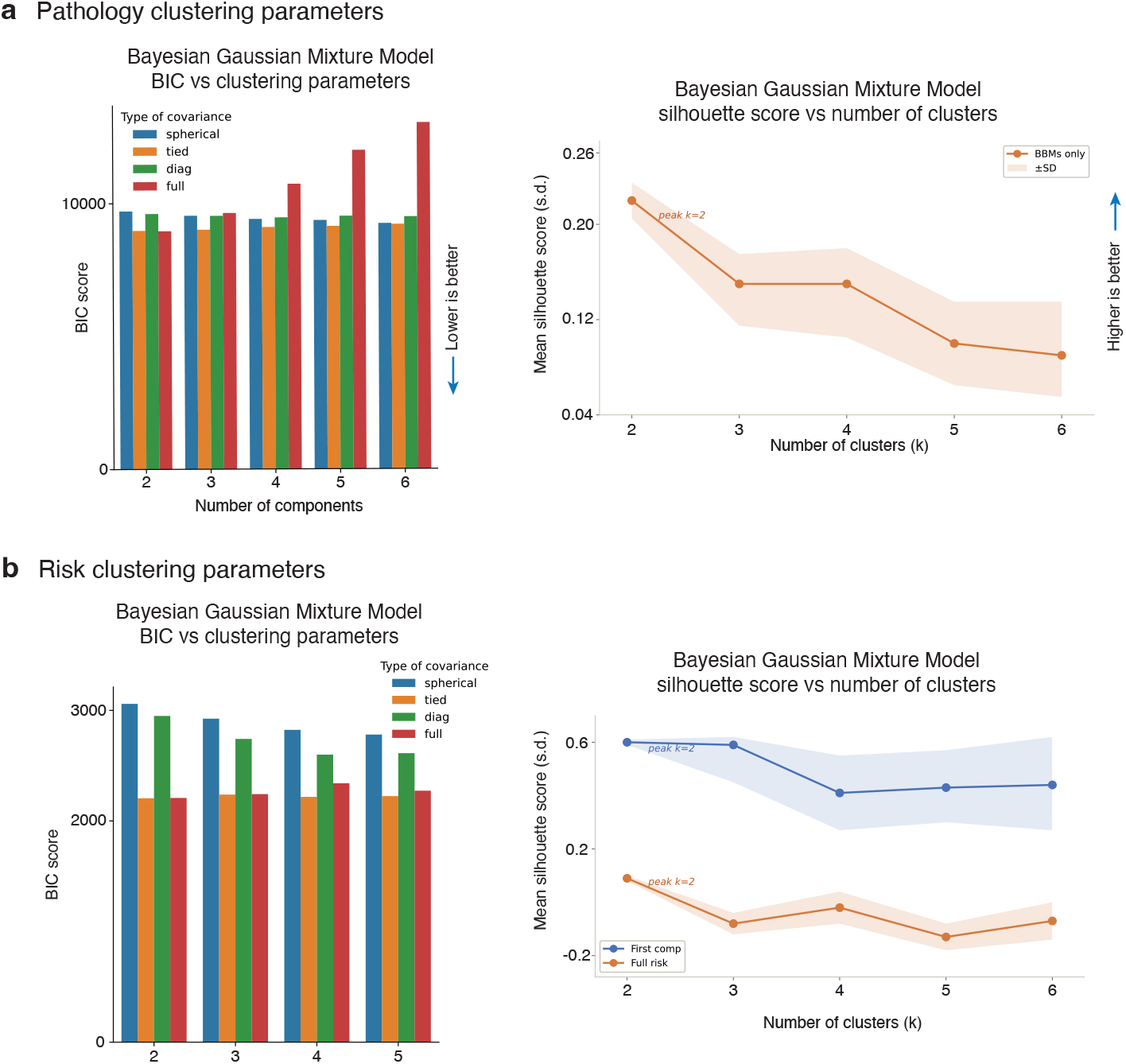
Clustering parameter selection for pathology and risk-based Bayesian Gaussian Mixture Models.a, Parameter optimization for the pathology clustering model. (Left) Bayesian Information Criterion (BIC) scores across covariance types (spherical, tied, diagonal, and full) and number of components (2–6). Lower BIC values indicate better model fit; best is full covariance with two components. (Right) Mean silhouette score (in standard deviations) as a function of the number of clusters (k = 2–6) for BBMs only, with shaded bands indicating ±1 s.d. The silhouette score peaks at k = 2, supporting a two-cluster solution as optimal for pathology-based stratification. b, Parameter optimization for the risk-based clustering model. (Left) BIC scores across covariance types and number of components (2–5), with tied covariance consistently achieving the lowest BIC. (Right) Mean silhouette scores for the first principal component (blue) and full risk score (orange) across k = 2–6 clusters, with shaded bands indicating ±1 s.d. Both measures peak at k = 2, corroborating the selection of two clusters as the optimal number for risk-based patient stratification.

