## Supplementary Materials for "Multivariate blood biomarkers capture resilience and resistance phenotypes across the Alzheimer’s disease spectrum"

#### Supplementary Figures

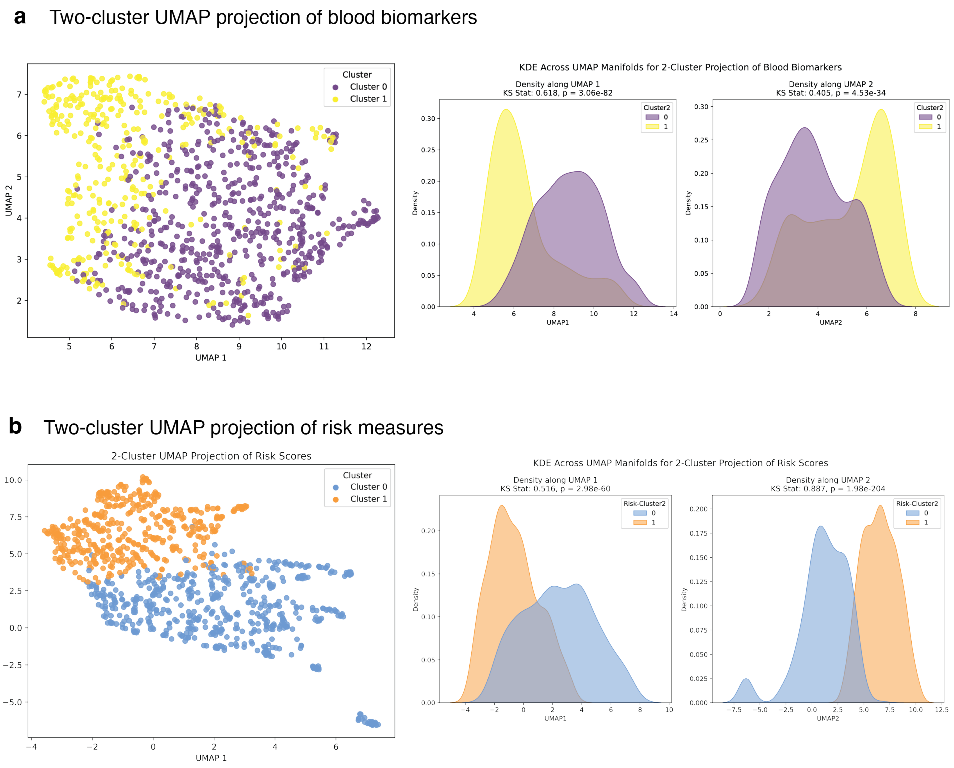


**Supplementary Fig. S1**: **UMAP clustering validity for blood biomarkers and risk measures. a,** Two-cluster UMAP projection of blood biomarkers. *Left*: UMAP embedding of participants coloured by cluster assignment (Cluster 0 (low), purple; Cluster 1 (high), yellow), demonstrating spatial separation of the two groups across the UMAP manifold. Right: Kernel density estimates (KDE) of cluster distributions along UMAP dimension 1 (KS statistic = 0.618, $p < 0.001$) and UMAP dimension 2 (KS statistic = 0.405, $p < 0.001$), confirming statistically significant separation between clusters. **b,** Two-cluster UMAP projection of risk measures. *Left*: UMAP embedding coloured by cluster assignment (Cluster 0 (low), blue; Cluster 1 (high), orange), illustrating distinct spatial organization of risk-based clusters. Right: KDE plots along UMAP dimension 1 (KS statistic = 0.516, $p < 0.001$) and UMAP dimension 2 (KS statistic = 0.887, $p < 0.001$), indicating highly significant distributional differences between the two risk clusters. KS: Kolmogorov–Smirnov.

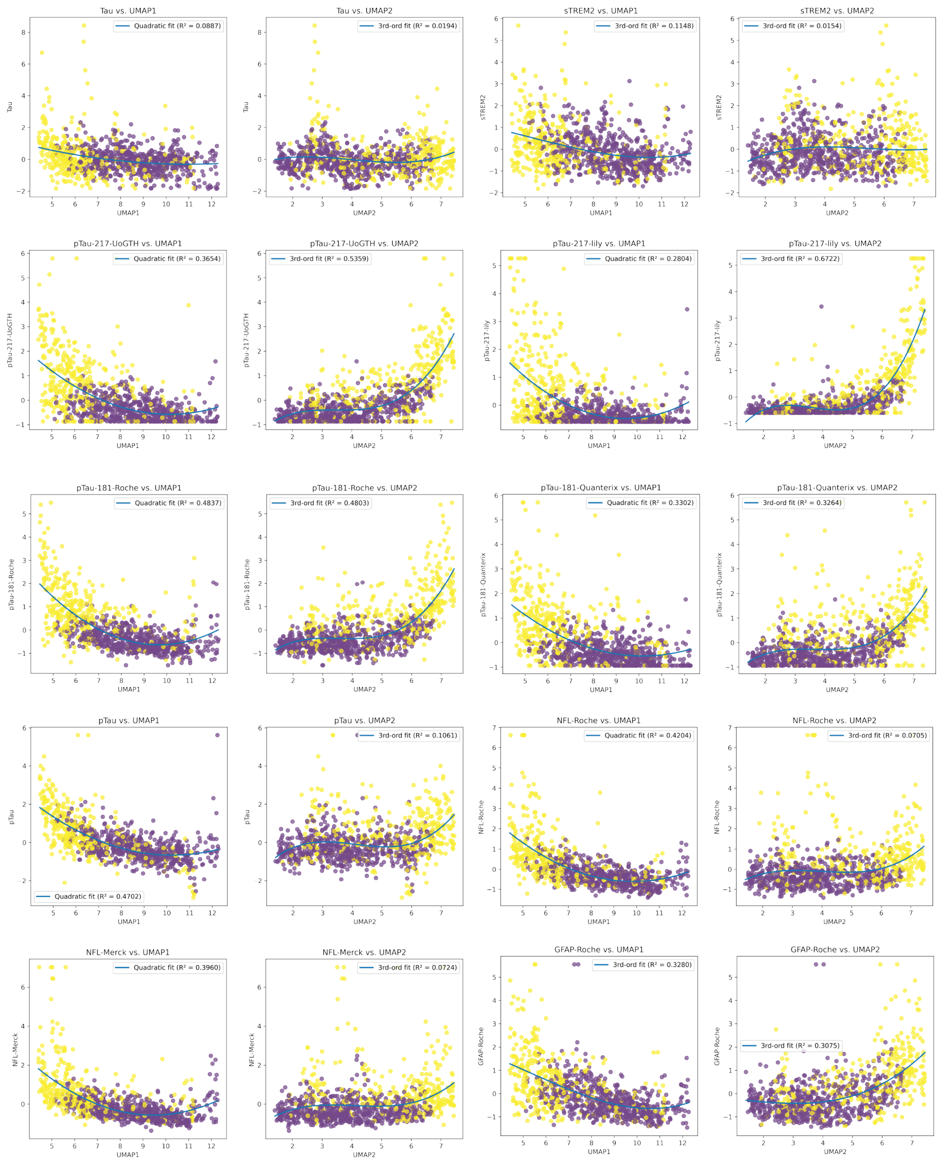


**Supplementary Fig. S2**: **Relationship between blood-based biomarkers and UMAP dimensions in the two-cluster blood biomarker projection**. Scatter plots depicting the association of each blood biomarker with UMAP dimension 1 (UMAP1) and UMAP dimension 2 (UMAP2) across participants assigned to Cluster 0 (low; purple) and Cluster 1 (high; yellow). Biomarkers shown include total Tau, sTREM2, p-tau217 (UoGTH and Lilly assays), p-tau181 (Roche and Quanterix assays), p-tau, NfL (Roche and Merck assays), and GFAP (Roche). Overlaid curves indicate the best-fitting polynomial regression (linear, quadratic or cubic, as indicated), with corresponding $R^2$ values reported in each panel title.

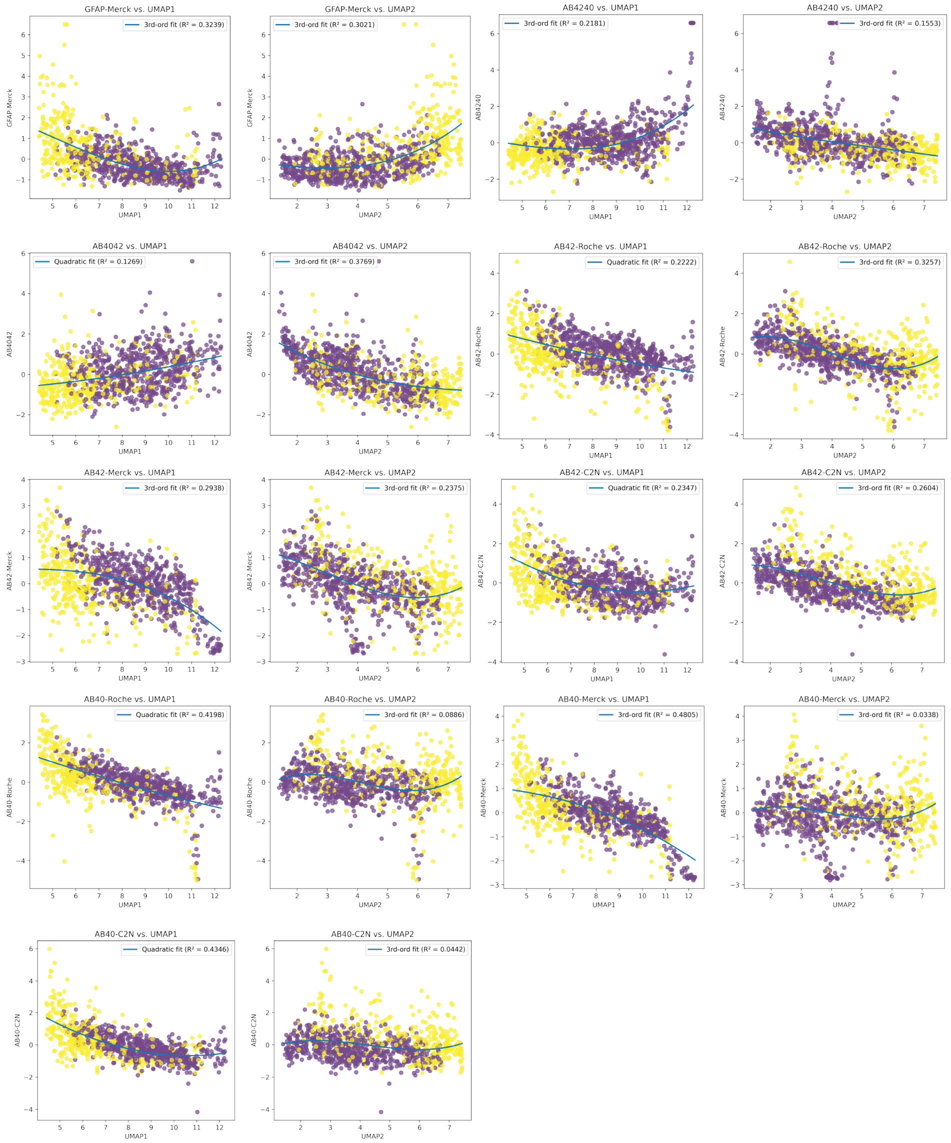


**Supplementary Fig. S3**: **Relationship between blood-based biomarkers and UMAP dimensions in the two-cluster blood biomarker projection (continued)**. Continuation of Supplementary Fig. S2. Biomarkers include GFAP (Merck), A$\beta$40/42 (Roche,Merck,and C2N assays), A$\beta$42 (Roche, Merck, and C2N assays), and A$\beta$40 (Roche, Merck, and C2N assays). Overlaid curves indicate the best-fitting polynomial regression (linear, quadratic or cubic, as indicated), with corresponding $R^2$ values reported in each panel title.

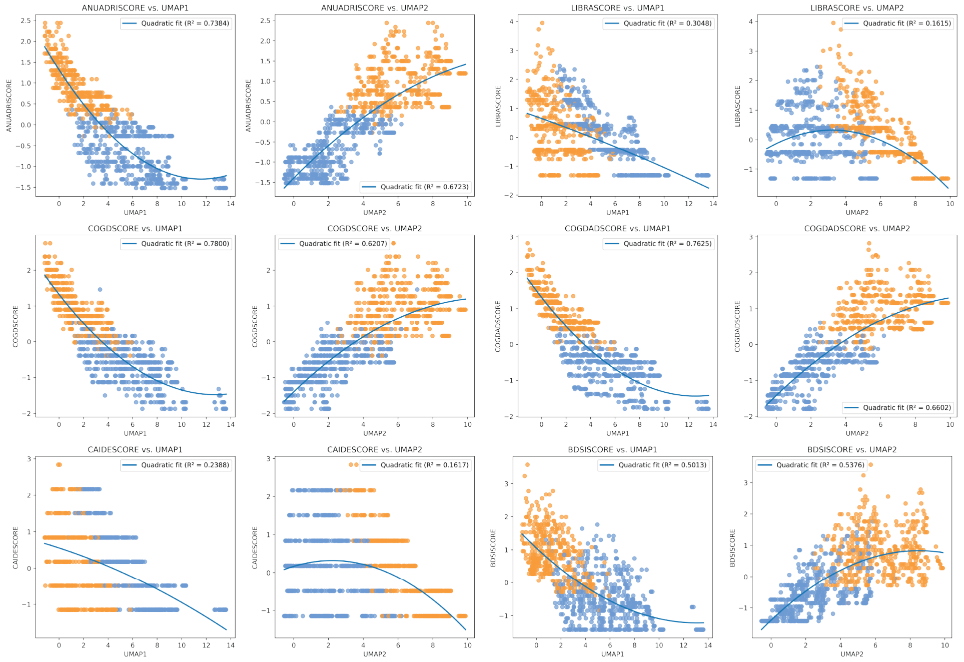


**Supplementary Fig. S4**: **Association of individual dementia risk scores with UMAP dimensions in the two-cluster risk measure projection.** Scatter plots illustrating the relationship between standardized risk scores and UMAP dimension 1 (UMAP1) and UMAP dimension 2 (UMAP2), with participants coloured by cluster membership (Cluster 0 (low), blue; Cluster 1 (high), orange). Scores depicted include ANUA-DRI, LIBRA, COGDrisk, COGDrisk-AD, CAIDE, and BDSI. Overlaid curves indicate the best-fitting polynomial regression (linear, quadratic or cubic, as indicated), with corresponding $R^2$ values reported in each panel title.

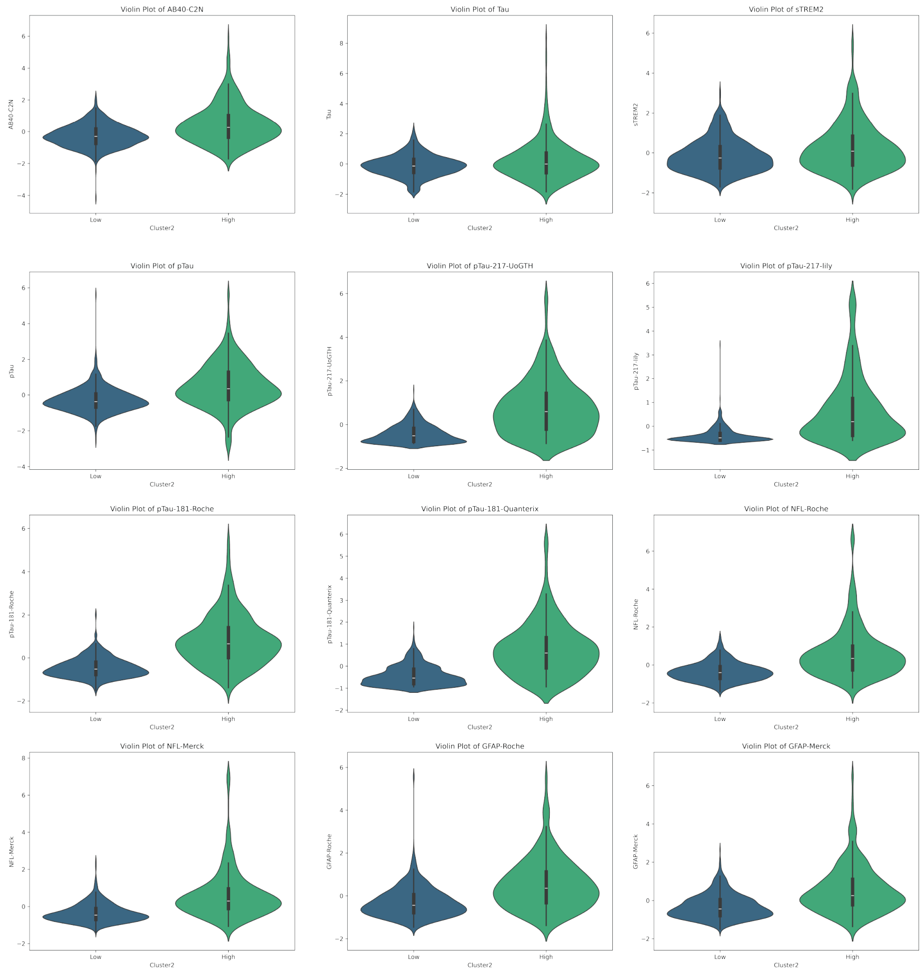


**Supplementary Fig. S5**: **Distribution of blood-based biomarkers across low- and high-pathology clusters.** Violin plots depicting the distribution of standardized blood biomarker values in participants assigned to the low-pathology (blue) and high-pathology (green) clusters. Biomarkers shown include A$\beta$40 (C2N), Tau, sTREM2, p-tau, p-tau217 (UoGTH), p-tau217 (Lilly), p-tau181 (Roche), p-tau181 (Quanterix), NfL (Roche), NfL (Merck), GFAP (Roche), and GFAP (Merck). The high-pathology cluster consistently exhibited elevated levels of p-tau, NfL, and GFAP relative to the low-pathology cluster. Internal horizontal lines denote median and interquartile range.

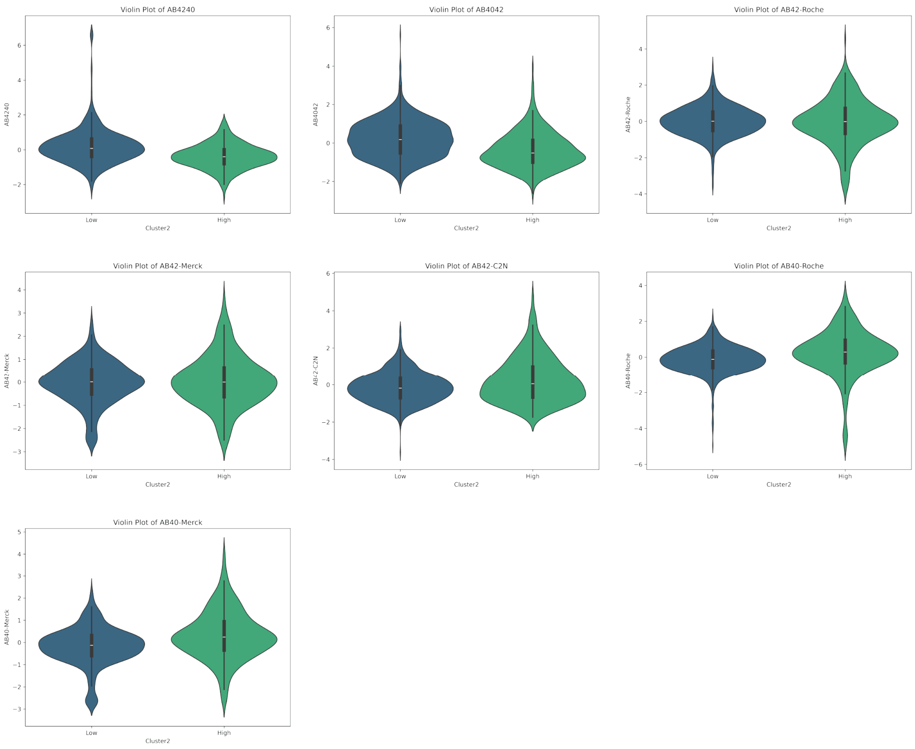


**Supplementary Fig. S6**: **Distribution of amyloid-$\beta$ species across low- and high-pathology clusters.** Violin plots showing the distribution of standardized plasma amyloid-$\beta$ biomarker concentrations in participants assigned to the low-pathology (blue) and high-pathology (green) clusters. Biomarkers include A$\beta$42/40, A$\beta$40/42, A$\beta$42-Roche, A$\beta$42-Merck, A$\beta$42-C2N, A$\beta$40-Roche, and A$\beta$40-Merck. Distinct distributional differences between clusters were observed across multiple amyloid species and assay platforms, with the high-pathology cluster generally exhibiting divergent amyloid profiles consistent with amyloid dysregulation. Internal horizontal lines represent the median and interquartile range.

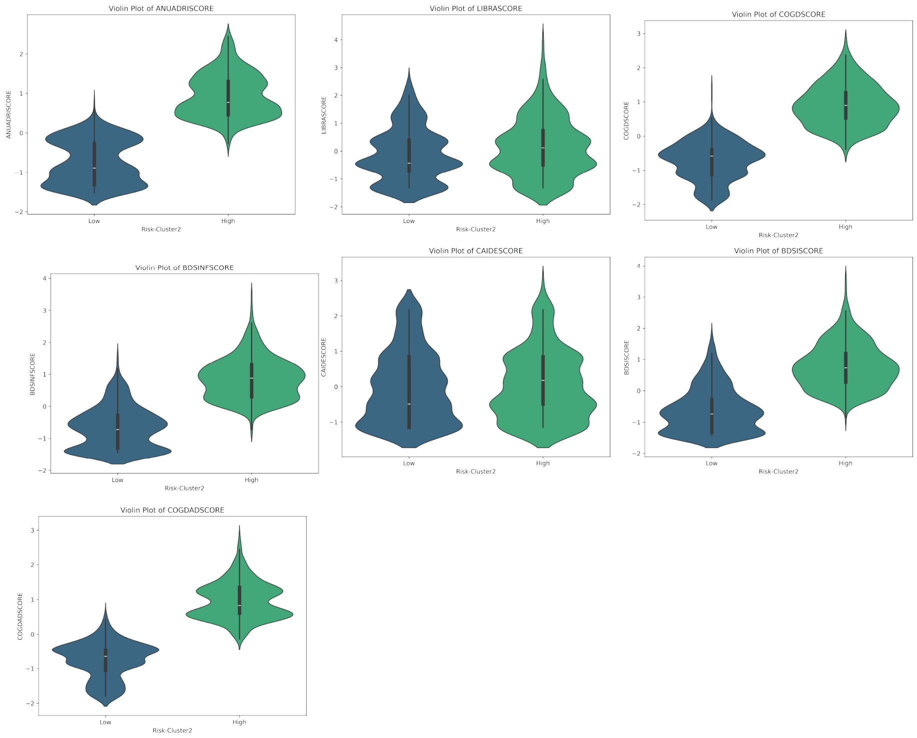


**Supplementary Fig. S7**: **Distribution of cognitive and functional risk scores across low- and high-risk clusters.** Violin plots illustrating the distribution of standardised cognitive and functional composite scores in participants assigned to the low-risk (blue) and high-risk (green) clusters. Scores depicted include ANU-ADRI, LIBRA, COGDrisk, BDSI-NF, CAIDE, BDSI, and COGDrisk-AD. Participants in the high-risk cluster demonstrated consistently elevated scores relative to the low-risk cluster, validating the clinical relevance of the risk-based two-cluster solution. Internal horizontal lines denote median and interquartile range.

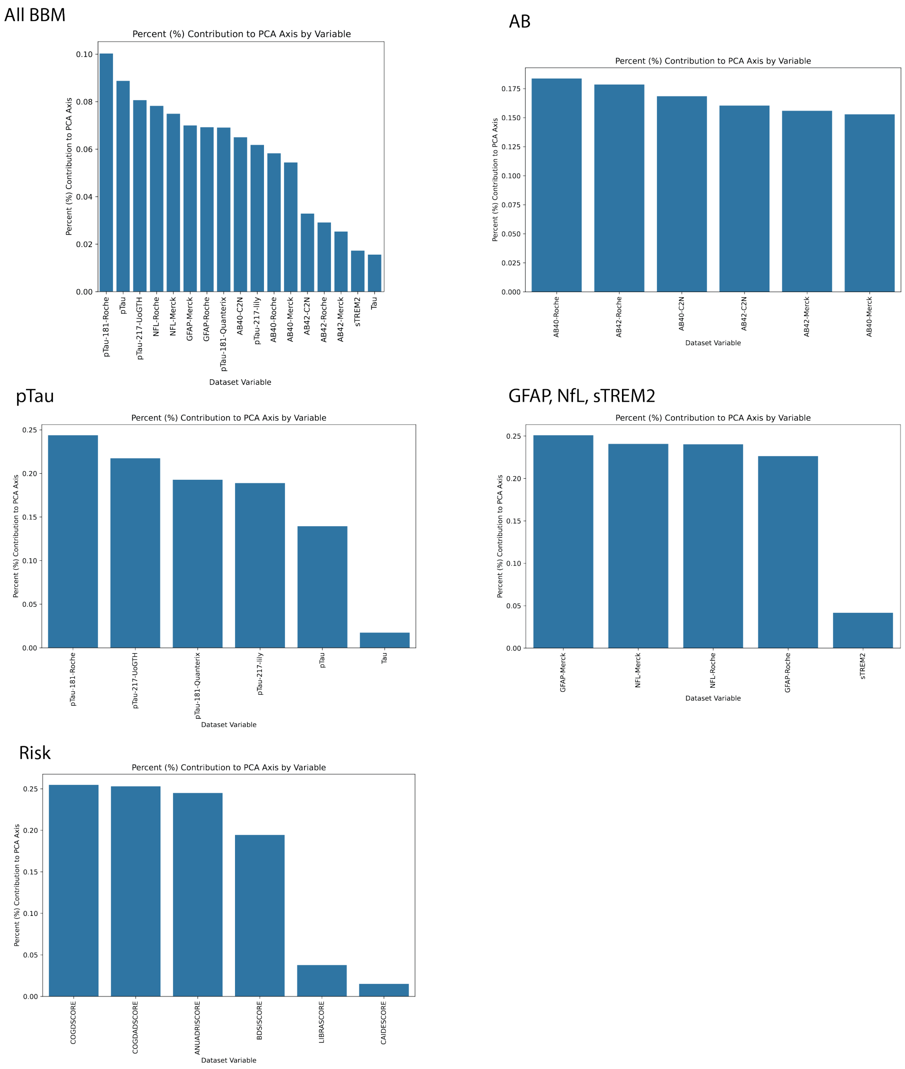


**Supplementary Fig. S8**: **PCA index loadings % contributions by feature, across indices.** Bar plots show the percent contribution of individual feature loading in the first PC of each derived PCA index (Method 8 from Figs. 2, 4.

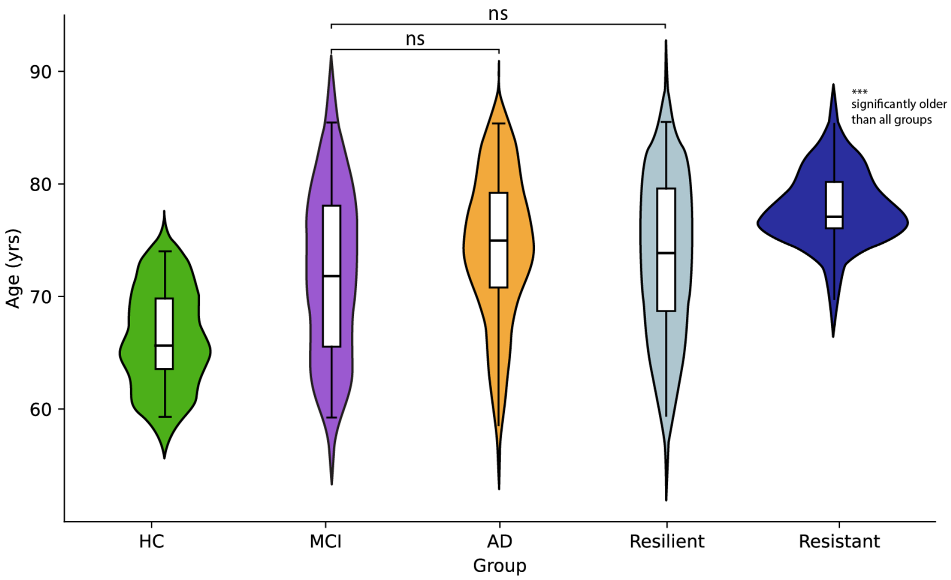


**Supplementary Fig. S9**: **Violin Plots of Age per Group.** Violin plots illustrating the distribution of age in participants assigned HC (green), MCI (purple), AD (orange), Resilient (light blue), Resistant (navy). Pairwise statistics revealed no significant differences between MCI or AD compared to Resilient group. Resistant group was significantly older than all groups. See Supplementary Table ST17 for full pairwise significance on age differences between groups.

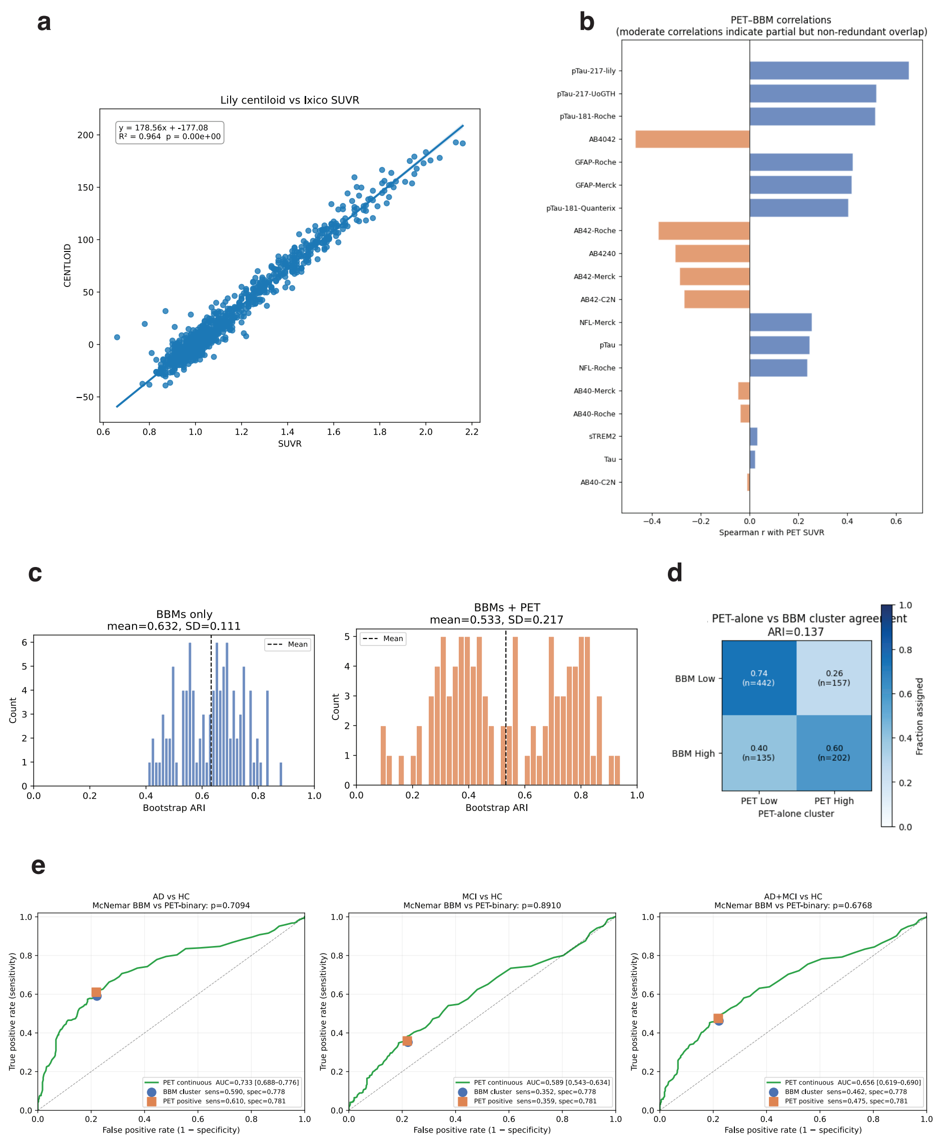


**Supplementary Fig. S10**: **Validation of PET quantification, blood biomarker–PET relationships, and comparison of PET-based and BBM-based clustering. a,** Scatter plot showing the correspondence between two PET amyloid quantification pipelines (Lily centiloid vs. Ixico SUVR) across all participants with available PET data. The strong linear relationship ($R^2 = 0.964$, $p < 0.001$) validates the interchangeability of the two pipelines for downstream analyses. **b,** Spearman correlations between individual blood biomarker measures (BBMs) and PET SUVR. Tau and neuroinflammatory markers (NfL, GFAP, sTREM2) show positive correlations, while A$\beta$ measures (orange) show negative correlations, consistent with the known inverse relationship between plasma A$\beta$42 and amyloid burden. Moderate correlation magnitudes indicate that BBMs capture partially overlapping but non-redundant information relative to PET. **c,** Bootstrap adjusted Rand index (ARI) distributions quantifying internal cluster stability for BBM-only (left; mean = 0.632, s.d. = 0.111) and BBM + PET (right; mean = 0.533, s.d. = 0.217) clustering solutions. Adding PET to the clustering reduces stability and introduces substantially greater variance, with a notably more dispersed and bimodal distribution, suggesting that PET and BBM features drive partially discordant cluster structure. **d,** Confusion matrix comparing cluster assignments derived independently from PET-alone versus BBM-alone (ARI = 0.137), confirming that the two modalities yield largely distinct groupings. **e,** Receiver operating characteristic (ROC) curves for classifying AD vs. HC (left), MCI vs. HC (middle), and AD + MCI vs. HC (right) using PET SUVR (continuous; green curve), BBM-derived cluster membership (blue point), and PET-positive binary threshold (orange point). McNemar tests indicate no significant difference in diagnostic performance between BBM-based and PET-binary classification across all comparisons (all $p > 0.67$).

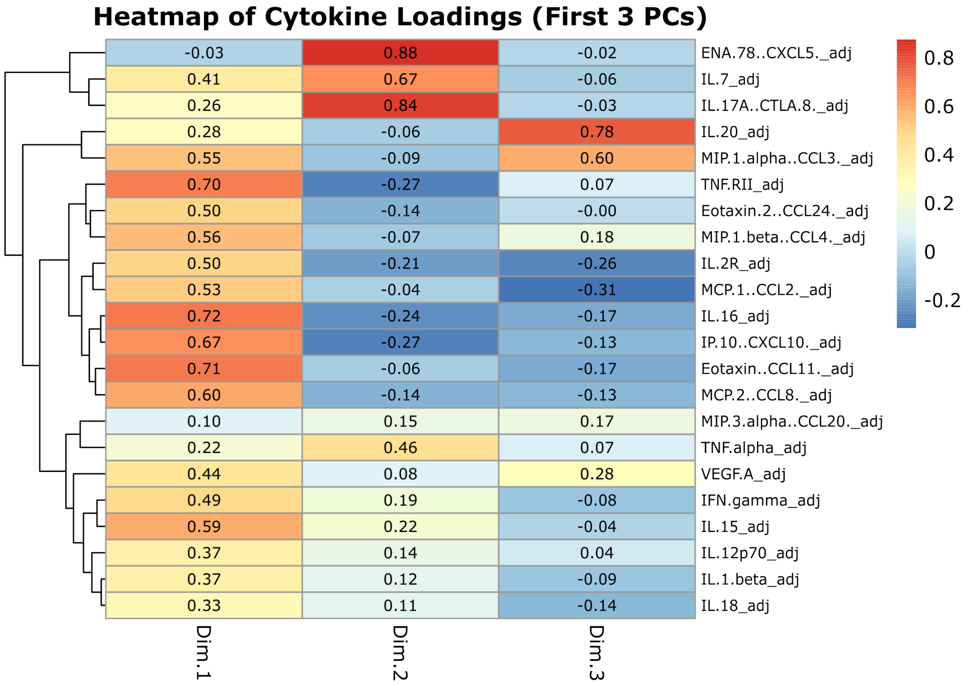


**Supplementary Fig. S11**: **Cytokine loadings on the first three principal components after varimax rotation.** Heatmap showing loadings of 24 adjusted serum cytokine measures on the first three principal components (Dim.1–Dim.3) derived from principal component analysis followed by varimax rotation. Cytokines (rows) are ordered by hierarchical clustering (dendrogram, left) based on similarity of their loading patterns across the three components. Cell color and value indicate the loading of each cytokine on each rotated component, with warm colors (red) denoting strong positive loadings and cool colors (blue) denoting negative loadings, as shown in the color scale. Dim.1 captures a broad axis of cytokine covariation (e.g., TNF.RII, IL-16, Eotaxin/CCL11), Dim.2 is dominated by ENA-78/CXCL5, IL-7, and IL-17A, and Dim.3 is characterized by high loadings for IL-20 and MIP-1$\alpha$/CCL3. Varimax rotation was applied to the top three unrotated components to improve interpretability by maximizing the variance of loadings within each component while preserving orthogonality.

### Supplementary Tables

**Supplementary Table ST1**: Permutation test of clustering quality for the pathology clusters. *Note.* The null distribution comprises 10,000 random label permutations (the observed value was excluded from the null). The 95% CI is the 2.5–97.5 percentile interval of the null distribution. $p$-values are one-sided (proportion of permutations $\geq$ the observed value) with a +1 correction; the minimum attainable value is 1/10,001 < 0.0001. ARI: adjusted Rand index.

| Metric | Observed | Null mean | Null 95% CI | $p$ |
|---|---|---|---|---|
| Silhouette | 0.208 | 0.062 | $[0.030, 0.100]$ | 0.0052 |
| ARI | 1.000 | 0.000 | $[-0.008, 0.012]$ | $< 0.0001$ |

**Supplementary Table ST2**: Permutation test of clustering quality for the risk clusters. *Note.* The null distribution comprises 10,000 random label permutations (the observed value was excluded from the null). The 95% CI is the 2.5–97.5 percentile interval of the null distribution. $p$-values are one-sided (proportion of permutations $\geq$ the observed value) with a +1 correction; the minimum attainable value is 1/10,001 < 0.0001. ARI: adjusted Rand index.

| Metric | Observed | Null mean | Null 95% CI | $p$ |
|---|---|---|---|---|
| Silhouette | 0.364 | 0.125 | $[0.117, 0.139]$ | $< 0.0001$ |
| ARI | 1.000 | 0.000 | $[-0.003, 0.006]$ | $< 0.0001$ |

**Supplementary Table ST3: Diagnostic criteria used for group classification.** *Note.* HC = healthy cognition; MCI = mild cognitive impairment; AD = Alzheimer's dementia; MMSE = Mini-Mental State Examination [1]; RAVLT = Rey Auditory Verbal Learning Test [2]; FAQ = Functional Activities Questionnaire [3]; NIA-AA = National Institute on Aging–Alzheimer's Association [4]. Criteria adapted from [5].

| Modality of criterion | HC ($N = 417$) | MCI ($N = 312$) | Mild AD ($N = 272$) |
|---|---|---|---|
| Alternative | — | Diagnosis of MCI meeting NIA-AA criteria and corroborated by medical records. | Diagnosis of probable AD meeting NIA-AA criteria and corroborated by medical records. |
| Cognition | MMSE of 26–30; and RAVLT delayed-recall performance within the normal range relative to age- and race-adjusted norms. | MMSE of 24–30; and RAVLT delayed recall at least 1 SD below the age- and race-adjusted mean. | MMSE of 20–24 (as low as 17 where warranted by clinical judgment); and RAVLT delayed recall at least 1 SD below the age- and race-adjusted mean. |
| Function | No functional decline evident on the FAQ (approximately 0–8) or informant report, in the investigator's judgment. | Minimal-to-mild functional impairment with independence largely preserved, based on the FAQ or informant report together with the investigator's judgment. | Functional decline with reduced independence, based on the FAQ or informant report together with the investigator's judgment. |

**Supplementary Table ST4**: Proteomic analysis results. Due to the size of this table, it is provided as a separate supplementary file: `Supplementary_Table_ST4.xlsx`

**Supplementary Table ST5**: Logistic regression with genomic predictors.

| Outcome | Term | $\beta$ | SE | $p$ | AUC |
|---|---|---|---|---|---|
| High Pathology and High Risk ($N = 169$) compared to Resistant ($N = 67$) | AD | 0.49 | 0.15 | <.001 | 0.62 |
| | AD no *APOE* | 0.29 | 0.15 | .05 | 0.57 |
| | ADRD | 0.57 | 0.15 | <.001 | 0.65 |
| | ADRD no *APOE* | 0.38 | 0.14 | .007 | 0.61 |
| | Education Attainment | −0.21 | 0.14 | .13 | 0.55 |
| | Cognition | −0.04 | 0.14 | .74 | 0.50 |
| | *APOE* ($\varepsilon 2$ and $\varepsilon 4$) | 0.45 | 0.17 | .007 | 0.63 |
| High pathology MCI ($N = 88$) compared to Resilient ($N = 73$) | AD | −0.004 | 0.16 | .97 | 0.52 |
| | AD no *APOE* | −0.04 | 0.16 | .77 | 0.52 |
| | ADRD | 0.09 | 0.16 | .59 | 0.52 |
| | ADRD no *APOE* | 0.03 | 0.15 | .84 | 0.52 |
| | Education Attainment | −0.26 | 0.15 | .08 | 0.58 |
| | Cognition | −0.15 | 0.16 | .36 | 0.53 |
| | *APOE* ($\varepsilon 2$ and $\varepsilon 4$) | 0.28 | 0.16 | .09 | 0.59 |
| High pathology AD ($N = 117$) compared to Resilient ($N = 73$) | AD | 0.21 | 0.15 | .16 | 0.55 |
| | AD no *APOE* | 0.20 | 0.15 | .19 | 0.55 |
| | ADRD | 0.22 | 0.15 | .15 | 0.56 |
| | ADRD no *APOE* | 0.09 | 0.15 | .53 | 0.53 |
| | Education Attainment | −0.21 | 0.15 | .15 | 0.56 |
| | Cognition | −0.04 | 0.14 | .77 | 0.51 |
| | *APOE* ($\varepsilon 2$ and $\varepsilon 4$) | 0.48 | 0.15 | .002 | 0.65 |

*Note.* Counts reflect number of cases with complete genomic data. Polygenic risk scores were adjusted for age, sex, and race. SE = standard error.

**Supplementary Table ST6**: Fisher exact test results for *APOE* variant frequency between groups within each comparison. *P* values were adjusted using the Benjamini–Hochberg method within each comparison, across the 6 variant tests.

| Comparison | Variant | $n_1$ | $n_2$ | OR | $p$ | $p_{\text{adj}}$ | Sig. |
|---|---|---|---|---|---|---|---|
| High Pathology and High Risk vs. Resistant | E2/E3 | 16 | 3 | 2.14 | .295 | .354 | ns |
| | E3/E3 | 102 | 59 | 0.35 | <.001 | .001 | ** |
| | E3/E4 | 76 | 15 | 2.49 | .005 | .014 | * |
| | E2/E4 | 5 | 2 | 0.96 | 1.000 | 1.000 | ns |
| | E4/E4 | 11 | 1 | 4.41 | .189 | .354 | ns |
| | E2/E2 | 0 | 1 | 0.00[a] | .278 | .354 | ns |
| High Pathology MCI vs. Resilient | E2/E3 | 11 | 12 | 0.76 | .657 | 1.000 | ns |
| | E3/E3 | 49 | 48 | 0.76 | .392 | 1.000 | ns |
| | E3/E4 | 39 | 24 | 1.60 | .168 | 1.000 | ns |
| | E2/E4 | 4 | 4 | 0.85 | 1.000 | 1.000 | ns |
| | E4/E4 | 4 | 3 | 1.14 | 1.000 | 1.000 | ns |
| | E2/E2 | 0 | 0 | –[b] | 1.000 | 1.000 | ns |
| High Pathology AD vs. Resilient | E2/E3 | 9 | 12 | 0.40 | .057 | .130 | ns |
| | E3/E3 | 65 | 48 | 0.63 | .086 | .130 | ns |
| | E3/E4 | 65 | 24 | 1.95 | .020 | .121 | ns |
| | E2/E4 | 3 | 4 | 0.42 | .263 | .315 | ns |
| | E4/E4 | 16 | 3 | 3.29 | .080 | .130 | ns |
| | E2/E2 | 0 | 0 | –[b] | 1.000 | 1.000 | ns |

*Note.* $n_1$ and $n_2$ give the number of variant carriers in group 1 and group 2, respectively. OR = Fisher's exact test odds ratio. Significance codes: * $p_{\text{adj}} \leq .05$; ** $p_{\text{adj}} \leq .01$; *** $p_{\text{adj}} \leq .001$; ns = not significant. [a] Boundary odds ratio estimate; no carriers observed in one of the two groups. [b] Odds ratio undefined; no carriers of this variant observed in either group, so this test is uninformative.

**Supplementary Table ST7**: Logistic regression with cytokine principal components as predictors. *Note.* $\chi^2$ denotes contribution to explaining variance based on ANOVA performed on the regression model. SE = standard error; OR = odds ratio; CI = 95% confidence interval; PC = Principal Component.

| Outcome | Term | Log Odds | SE | Wald $z$ | $p$ | OR | OR CI | $\chi^2$ | $\chi^2$ $p$ |
|---|---|---|---|---|---|---|---|---|---|
| Resistant ($N = 79$) compared to High Pathology and High Risk ($N = 203$) | Intercept | −1.052 | 1.599 | −0.658 | .511 | 0.35 | [0.01, 7.98] | – | – |
| | Age | 0 | 0 | 1.242 | .214 | 1.00 | [1.00, 1.00] | 0.75 | .387 |
| | Sex | 0.115 | 0.270 | 0.426 | .670 | 1.12 | [0.66, 1.91] | 0.14 | .706 |
| | PC1 | 0.127 | 0.064 | 1.994 | .046 | 1.14 | [1.00, 1.29] | 3.35 | .067 |
| | PC2 | 0.173 | 0.089 | 1.942 | .052 | 1.19 | [1.00, 1.42] | 3.84 | .050 |
| | PC3 | −0.037 | 0.114 | −0.324 | .746 | 0.96 | [0.77, 1.21] | 0.10 | .747 |
| Resilient ($N = 86$) compared to High Pathology MCI ($N = 103$) | Intercept | −0.208 | 0.919 | −0.227 | .821 | 0.81 | [0.13, 4.95] | – | – |
| | Age | 0 | 0 | 0.277 | .782 | 1.00 | [1.00, 1.00] | 0.04 | .849 |
| | Sex | 0.341 | 0.301 | 1.133 | .257 | 1.41 | [0.78, 2.55] | 0.96 | .326 |
| | PC1 | 0.046 | 0.068 | 0.672 | .502 | 1.05 | [0.92, 1.20] | 0.36 | .550 |
| | PC2 | 0.043 | 0.091 | 0.473 | .636 | 1.04 | [0.87, 1.25] | 0.15 | .699 |
| | PC3 | −0.178 | 0.122 | −1.458 | .145 | 0.84 | [0.65, 1.06] | 2.17 | .141 |
| Resilient ($N = 86$) compared to High Pathology AD ($N = 153$) | Intercept | −1.002 | 0.914 | −1.096 | .273 | 0.37 | [0.06, 2.20] | – | – |
| | Age | 0 | 0 | 1.610 | .107 | 1.00 | [1.00, 1.00] | 1.58 | .209 |
| | Sex | 0.160 | 0.284 | 0.562 | .574 | 1.17 | [0.67, 2.05] | 0.41 | .523 |
| | PC1 | 0.183 | 0.063 | 2.915 | .004 | 1.20 | [1.06, 1.36] | 8.97 | .003 |
| | PC2 | 0.253 | 0.088 | 2.888 | .004 | 1.29 | [1.09, 1.54] | 7.94 | .005 |
| | PC3 | −0.233 | 0.115 | −2.032 | .042 | 0.79 | [0.63, 0.99] | 4.13 | .042 |

**Supplementary Table ST8**: Protein biomarkers, assays, and the vendor providing each result.

| Biomarker Category | Biomarker Name | Vendor Providing Result |
|---|---|---|
| Amyloid Beta (AB) | A$\beta$40 | C2N<br>Merck<br>Roche |
| | A$\beta$42 | C2N<br>Merck<br>Roche |
| | A$\beta$40/42 | C2N<br>Merck |
| Phosphorylated Tau (Tau) | total p-tau | Clinical dataset provider |
| | p-tau181 | Quanterix<br>Roche |
| | p-tau217 | University of Gothenburg<br>Lilly |
| | total Tau | Quanterix |
| Other neuropathology markers | Glial Fibrillary Acidic Protein (GFAP) | Merck<br>Roche Diagnostics |
| | Neurofilament light chain (NfL) | Merck<br>Roche Diagnostics |
| | Soluble Triggering Receptor Expressed on Myeloid cells-2 | Clinical dataset provider |

**Supplementary Table ST9**: Upper-bound thresholds applied for outlier winsorization by biomarker assay. *Note.* Values above each threshold were set to the threshold value (winsorized), prior to a final outlier removal step using a z-score cutoff of 8 s.d. applied to all biomarkers. In some cases, if a minimum value was given like "<LLOQ", "<LOD", "<0.1", "<0.01", then these were imputed using the lowest numerical value in the cohort sample.)

| Biomarker (assay) | Threshold |
|---|---|
| p-tau217 (Lilly) | 20 |
| p-tau217 (University of Gothenburg) | 0.017 |
| AB40/42 ratio (C2N) | 0.17 |
| Tau (Quanterix) | 0.02 |
| p-tau (clinical dataset provider) | 0.15 |
| NfL (Merck) | 0.2 |
| NfL (Roche) | 0.02 |
| GFAP (Merck) | 1.0 |
| GFAP (Roche) | 0.6 |
| p-tau181 (Quanterix) | 0.1 |

**Supplementary Table ST10**: PC weights used to compute the composite dementia risk index. Seven standardized risk scores (ANU-ADRI, COGDrisk-AD, COGDrisk, BDSI-NF, BDSI, CAIDE, LIBRA) were submitted to PCA after mean-centring. PC1 (64.8 % variance explained) reflects the shared dementia-risk signal across all instruments. PC2 (24.0 % variance explained) captures a contrast between lifestyle-based indices (CAIDE, LIBRA; positive loadings) and the clinical scoring instruments (negative loadings), and is not used in the classification model. The composite index is computed from PC1 only: Index $= \sum_j w_j^{(1)}(x_j - \bar{x}_j)$, where $w_j^{(1)}$ is the PC1 coefficient.

| Risk score | Feature mean | PC1 loading | PC1 coefficient | PC2 loading | PC2 coefficient |
|---|---|---|---|---|---|
| ANU-ADRI | -0.1149 | 0.4579 | 0.2184 | -0.0820 | -0.0643 |
| COGDrisk-AD | -0.1387 | 0.4583 | 0.2186 | -0.0751 | -0.0589 |
| COGDrisk | -0.1170 | 0.4625 | 0.2206 | -0.0291 | -0.0228 |
| BDSI-NF | -0.1678 | 0.4335 | 0.2068 | -0.1080 | -0.0847 |
| BDSI | -0.2523 | 0.3862 | 0.1842 | -0.0647 | -0.0507 |
| CAIDE | -0.0084 | 0.0774 | 0.0369 | 0.7353 | 0.5765 |
| LIBRA | -0.0135 | 0.1529 | 0.0730 | 0.6560 | 0.5143 |
| % Variance explained | | 64.8 % | | 24.0 % | |

*Note.* Loadings are unit-normalized eigenvector components; coefficients equal the loading scaled by $\sqrt{\lambda_k}$ (square root of the $k$-th eigenvalue: $\lambda_1 = 4.395$, $\lambda_2 = 1.627$). CAIDE and LIBRA load weakly on PC1 relative to the clinical instruments, reflecting their partial independence from the shared clinical risk signal.

**Supplementary Table ST11**: Logistic regression model using the composite dementia risk index (PC1 of seven standardized risk scores) as a single predictor. Reported for completeness; the composite index does not outperform the recommended single-score models (ANU-ADRI, COGDrisk-AD; Supplementary Table ST12). Performance metrics are means ± s.d. across cross-validation folds.

| Predictor | $\beta$ | 95 % CI lower | 95 % CI upper | Intercept | Youden threshold | Bal. acc. mean ± SD |
|---|---|---|---|---|---|---|
| Risk PC1 | 5.984 | 5.473 | 6.537 | -0.973 | 0.418 | 0.961 ± 0.007 |

*Note.* AUC: 0.991 ± 0.003. The predicted log-odds is $\hat{y} = -0.973 + 5.984\,x$, where $x$ is the PC1 composite risk index (Supplementary Table ST10). Classification is positive when $\hat{p} = \sigma(\hat{y}) \geq 0.418$.

**Supplementary Table ST12**: Logistic regression models using individual dementia risk scores as single predictors. Models are ordered by cross-validated balanced accuracy; ANU-ADRI and COGDrisk-AD are the recommended models (highest balanced accuracy and AUC). Each model uses a single standardized risk score; the intercept and Youden's $J$-derived probability threshold are reported per model. Performance metrics are means ± s.d. across cross-validation folds. CI = 95 % bootstrap confidence interval.

| Risk score | $\beta$ | 95 % CI lower | 95 % CI upper | Intercept | Youden threshold | Bal. acc. mean ± SD |
|---|---|---|---|---|---|---|
| *Recommended* | | | | | | |
| ANU-ADRI | 7.219 | 6.670 | 7.778 | -1.284 | 0.452 | 0.977 ± 0.013 |
| COGDrisk-AD | 6.517 | 6.147 | 6.830 | -0.867 | 0.370 | 0.971 ± 0.006 |
| *Other evaluated models* | | | | | | |
| COGDrisk | 5.536 | 4.995 | 6.250 | -0.597 | 0.552 | 0.943 ± 0.013 |
| BDSI-NF | 3.548 | 3.189 | 3.958 | -0.600 | 0.357 | 0.912 ± 0.010 |
| BDSI | 2.500 | 2.261 | 2.812 | -0.401 | 0.296 | 0.858 ± 0.010 |

*Note.* AUC (mean ± SD): ANU-ADRI 0.995 ± 0.003; COGDrisk-AD 0.997 ± 0.002; COGDrisk 0.985 ± 0.004; BDSI-NF 0.955 ± 0.006; BDSI 0.914 ± 0.012. The predicted log-odds is $\hat{y} = \beta_0 + \beta\, x$, where $x$ is the standardized risk score. Classification is positive when $\hat{p} = \sigma(\hat{y}) \geq \tau_{\text{Youden}}$. See Supplementary Table ST18 for standardization parameters.

**Supplementary Table ST13**: PC1 weights used to compute each biomarker composite index. Each vendor-specific assay was centred by subtracting its training-set mean prior to projection. The composite index for a given sample is obtained as Index $= \sum_j w_j(x_j - \bar{x}_j)$, where $w_j$ is the PC1 coefficient and $\bar{x}_j$ is the feature mean listed below. Any subset of available assays may be used; the index remains comparable across cohorts provided at least one assay per biomarker is available.

| Biomarker | Assay | Feature mean | PC1 loading | PC1 coefficient |
|---|---|---|---|---|
| A$\beta$40 (*73.6 % var.*) | C2N | 1.135e-04 | 0.5649 | 0.3835 |
| | Merck | 3.897e-03 | 0.5586 | 0.3792 |
| | Roche | 0.0 | 0.6073 | 0.4123 |
| p-tau217 (*85.6 % var.*) | UoGTH | 0.0 | 0.7117 | 0.5467 |
| | Lilly | -3.873e-03 | 0.7025 | 0.5397 |
| p-tau181 (*86.9 % var.*) | Quanterix | 7.952e-04 | 0.7043 | 0.5361 |
| | Roche | 0.0 | 0.7099 | 0.5404 |
| NfL (*94.4 % var.*) | Merck | -2.061e-03 | 0.7053 | 0.5147 |
| | Roche | 0.0 | 0.7090 | 0.5174 |

*Note.* PC1 loadings are the unit-normalised eigenvector components; PC1 coefficients equal the loading scaled by $\sqrt{\lambda_1}$ (square root of the first eigenvalue) and are the values applied directly to mean-centred assay values to obtain the composite index score. Variance explained refers to the proportion of total variance captured by PC1 within each biomarker panel.

**Supplementary Table ST14**: Logistic regression coefficients for the three classification models. Predictors are either PCA-derived composite indices (Supplementary Table ST13) or individual vendor assays (best-single model). All models share an intercept term; the probability threshold was selected via Youden's $J$ statistic to maximise cross-validated balanced accuracy. Performance metrics are means ± s.d. across cross-validation folds. CI = 95 % bootstrap confidence interval.

| Model | Predictor | $\beta$ | 95 % CI lower | 95 % CI upper | Intercept |
|---|---|---|---|---|---|
| **A. 3-index model** (BA: 0.860 ± 0.026; AUC: 0.920 ± 0.024; Youden threshold: 0.434) | | | | | |
| | NfL index | 0.9620 | 0.7565 | 1.1924 | |
| | p-tau181 index | 1.2039 | 0.9220 | 1.5228 | −0.533 |
| | p-tau217 index | 0.8519 | 0.5040 | 1.1483 | |
| **B. 4-index model** (BA: 0.863 ± 0.032; AUC: 0.919 ± 0.026; Youden threshold: 0.452) | | | | | |
| | A$\beta$40 index | 0.1831 | 0.0599 | 0.3686 | |
| | NfL index | 0.8899 | 0.6932 | 1.1200 | −0.547 |
| | p-tau181 index | 1.1624 | 0.8593 | 1.4913 | |
| | p-tau217 index | 0.8775 | 0.5282 | 1.1699 | |
| **C. Best-single-vendor model** (BA: 0.846 ± 0.031; AUC: 0.896 ± 0.031; Youden threshold: 0.4[illegible]) | | | | | |
| | A$\beta$40 (Roche) | -0.0219 | -0.1851 | 0.2220 | |
| | NfL (Roche) | 1.1422 | 0.9185 | 1.3866 | −0.57[illegible] |
| | p-tau181 (Roche) | 1.4382 | 1.1180 | 1.7036 | |
| | p-tau217 (UoGTH) | 0.8395 | 0.5858 | 1.0851 | |

*Note.* The predicted log-odds for a new observation is $\hat{y} = \beta_0 + \sum_j \beta_j x_j$, where $\beta_0$ is the intercept and $x_j$ are the composite index scores (Panels A–B) or standardized individual assay values (Panel C). Classification is positive when $\hat{p} = \sigma(\hat{y}) \geq \tau_{\text{Youden}}$. See Supplementary Table ST18 for standardization parameters. BA: balanced accuracy.

**Supplementary Table ST15**: Variables included in each descriptive PCA composite index. Each index was derived from PC1 of its respective feature set after standardization. The cognition index was computed following $k$-nearest-neighbour imputation to recover missing observations across its constituent measures. Not described here explicitly is the "All pathology" index from the main text; this particular index was comprised of all features from the p-tau, A$\beta$, and "Glial/neurodegenerative pathology" indices, combined.

| Index | Features |
|---|---|
| Cognition | AVL Total Learning (AVL0235C)<br>DANA Guessing — Mean Throughput<br>DANA Procedural Reaction Time — Mean Throughput<br>DANA Simple Reaction Time — Mean Throughput<br>DCR Summary Score<br>Mean Image Description Score<br>Mean Image Description Volition<br>Mean Object Recall Score<br>Pathfinding — Time to Complete<br>SDMT Performance<br>Spiral Performance<br>Trails B Performance |
| Glial/neurodegenerative pathology | GFAP (Merck)<br>GFAP (Roche)<br>NfL (Merck)<br>NfL (Roche)<br>sTREM2 |
| Dementia risk | ANU-ADRI<br>BDSI<br>CAIDE<br>COGDrisk-AD<br>COGDrisk<br>LIBRA |
| Amyloid-$\beta$ | A$\beta$40 (C2N)<br>A$\beta$40 (Merck)<br>A$\beta$40 (Roche)<br>A$\beta$42 (C2N)<br>A$\beta$42 (Merck)<br>A$\beta$42 (Roche) |
| p-tau / Tau | Tau<br>p-tau<br>p-tau181 (Quanterix)<br>p-tau181 (Roche)<br>p-tau217 (UoGTH)<br>p-tau217 (Lilly) |

*Note.* These composite indices are descriptive and are distinct from the vendor-harmonization PCA indices used in the classification models (see Supplementary Table ST13). All features were mean-centred prior to PCA.

**Supplementary Table ST16**: Comparison of unified pathology axis scores between blood based biomarker (BBM) defined and PET defined phenotype groups. PCA index composition described in Supplementary Table ST15.

| Group | Test | Mean | PET Mean | $p$-value | Sig. | Cohen's $d$ | Effect Size | Direction |
|---|---|---|---|---|---|---|---|---|
| Resilient | A$\beta$ BBM PCA | 0.62 | -0.50 | $1.84 \times 10^{-9}$ | *** | 1.02 | large | positive |
| | All BBM PCA | 0.59 | -0.37 | $4.29 \times 10^{-12}$ | *** | 1.20 | large | positive |
| | Glial/NDG BBM PCA | 0.25 | -0.29 | $2.92 \times 10^{-4}$ | *** | 0.68 | medium | positive |
| | Risk PCA | 0.15 | -0.32 | 0.005 | ** | 0.51 | medium | positive |
| | p-tau BBM PCA | 0.45 | -0.17 | $5.37 \times 10^{-9}$ | *** | 0.98 | large | positive |
| Resistant | A$\beta$ BBM PCA | 0.14 | 1.25 | $7.01 \times 10^{-4}$ | *** | 1.06 | large | negative |
| | All BBM PCA | -0.28 | 0.85 | $3.39 \times 10^{-6}$ | *** | 1.60 | large | negative |
| | Glial/NDG BBM PCA | -0.26 | 0.48 | $1.50 \times 10^{-4}$ | *** | 0.98 | large | negative |
| | Risk PCA | 0.69 | 0.94 | 0.054 | ns | 0.50 | medium | negative |
| | p-tau PCA | -0.45 | 0.33 | $1.73 \times 10^{-5}$ | *** | 1.48 | large | negative |

**Supplementary Table ST17**: Pairwise significance comparisons of age between diagnostic groups.

| Group 1 | Group 2 | $n_1$ | $n_2$ | $p_{\text{adj}}$ |
|---|---|---|---|---|
| HC | MCI | 244 | 308 | $< .001$ |
| HC | AD | 244 | 266 | $< .001$ |
| MCI | AD | 308 | 266 | $< .001$ |
| HC | Resilient | 244 | 91 | $< .001$ |
| MCI | Resilient | 308 | 91 | .104 |
| AD | Resilient | 266 | 91 | .133 |
| HC | Resistant | 244 | 81 | $< .001$ |
| MCI | Resistant | 308 | 81 | $< .001$ |
| AD | Resistant | 266 | 81 | $< .001$ |
| Resilient | Resistant | 91 | 81 | $< .001$ |

Note. Values below .001 are reported as $< .001$. All $p$ values below .001 reached significance at the $p < .001$ threshold.

**Supplementary Table ST18**: Summary statistics of biomarker measures by source and test.

| Source | Test | Mean | SD | Min | Max |
|---|---|---|---|---|---|
| C2N | AB40-C2N | 0.501 | 0.0980 | 0.0930 | 1.09 |
| | AB4042-C2N | 0.0971 | 0.00984 | 0.0714 | 0.152 |
| | AB42-C2N | 0.0484 | 0.00946 | 0.0142 | 0.0942 |
| Merck | AB40-Merck | 0.109 | 0.0386 | 0.00028 | 0.266 |
| | AB4042-Merck | 0.0538 | 0.0161 | 0.0106 | 0.160 |
| | AB42-Merck | 0.00559 | 0.00199 | 0.000211 | 0.0129 |
| | GFAP-Merck | 0.180 | 0.106 | 0.0191 | 0.870 |
| | NfL-Merck | 0.0312 | 0.0205 | 0.00299 | 0.176 |
| | sTREM2 | 3.49 | 1.61 | 0.573 | 12.6 |
| Quanterix | Tau | 0.00208 | 0.00108 | 0.000076 | 0.0112 |
| | p-tau181-Quanterix | 0.0195 | 0.0114 | 0.00884 | 0.0845 |
| Roche | AB40-Roche | 0.297 | 0.0564 | 0.015 | 0.491 |
| | AB42-Roche | 0.0353 | 0.00870 | 0.00225 | 0.0749 |
| | GFAP-Roche | 0.113 | 0.0625 | 0.0214 | 0.459 |
| | NfL-Roche | 0.00347 | 0.00227 | 0.000256 | 0.0185 |
| | p-tau | 0.0211 | 0.00682 | 0.00131 | 0.0594 |
| | p-tau181-Roche | 0.00108 | 0.000570 | 0.000221 | 0.00421 |
| UoGTH | p-tau217-UoGTH | 0.00304 | 0.00213 | 0.0012 | 0.0154 |
| lily | p-tau217-lily | 3.11 | 2.58 | 1.57 | 16.7 |
